# Sex-specific genetic determinants of right ventricular structure and function

**DOI:** 10.1101/2024.02.06.23300256

**Authors:** Lars Harbaum, Jan K Hennigs, Julian Pott, Jonna Ostermann, Christoph R Sinning, Arunashis Sau, Ewa Sieliwonczyk, Fu Siong Ng, Christopher J Rhodes, Khodr Tello, Hans Klose, Stefan Gräf, Martin R Wilkins

**Affiliations:** Division of Respiratory Medicine, Department of Internal Medicine II, University Medical Centre Hamburg-Eppendorf, Hamburg, Germany; National Heart and Lung Institute, Faculty of Medicine, Imperial College London, London, United Kingdom; Department of Cardiology, University Heart and Vascular Centre Hamburg, University Medical Centre Hamburg-Eppendorf, Member of the German Center for Cardiovascular Research (DZHK), Partner Site Hamburg/Kiel/Luebeck, Hamburg, Germany; Department of Cardiology, Imperial College Healthcare NHS Trust, London, United Kingdom; MRC Laboratory of Medical Sciences, Imperial College London, London, United Kingdom; University of Antwerp and Antwerp University Hospital, Antwerp, Belgium; Department of Cardiology, Chelsea and Westminster Hospital NHS Foundation Trust, London, United Kingdom; Department of Internal Medicine, Justus-Liebig-University Giessen, Universities of Giessen and Marburg Lung Center, Member of the German Center for Lung Research (DZL), Giessen, Germany; NIHR BioResource for Translational Research - Rare Diseases, Department of Haemotology, University of Cambridge, Cambridge Biomedical Campus, Cambridge, United Kingdom; Department of Medicine, School of Clinical Medicine, University of Cambridge, Cambridge Biomedical Campus, Cambridge, United Kingdom

**Author notes:** Correspondence to: Lars Harbaum, MD, Division of Respiratory Medicine, Department of Internal Medicine II, University Medical Centre Hamburg-Eppendorf, Martinistraße 52, 20246 Hamburg, Germany.

## Abstract

**Background:** While sex differences in right heart phenotypes have been observed, the molecular drivers remain unknown. We used common genetic variation to provide biological insights into sex differences in the structure and function of the right ventricle (RV).

**Methods:** RV phenotypes were obtained from cardiac magnetic resonance imaging in 18,156 women and 16,171 men from the UK Biobank, based on a deep-learning approach, including end-diastolic, end-systolic, and stroke volumes, as well as ejection fraction. Observational analyses and sex-stratified genome-wide association studies were performed. Candidate female-specific loci were evaluated against invasively measured hemodynamics in 479 female patients with idiopathic or heritable pulmonary arterial hypertension (PAH), recruited to the UK National Institute for Health Research BioResource Rare Diseases study.

**Results:** Sex was associated with differences in RV volumes and ejection fraction in models adjusting for left heart counterparts and lung function. Six genome-wide significant loci (13%) revealed heterogeneity of allelic effects between women and men. These included two sex-specific candidate loci present in women only; namely, a locus for RV ejection fraction in *BMPR1A* and a locus for RV end-systolic volume near *DMRT2*. Epigenetic data indicate that variation at the *BMPR1A* locus likely alters transcriptional regulation in RV tissue. In female patients with PAH, a variant located in the promoter of *BMPR1A* was significantly associated with cardiac index (effect size 0.16 l/min/m^2^), despite similar RV afterload among genotypic groups.

**Conclusions:** We report sex-specific genetic loci for RV structure and function. *BMPR1A* has emerged as a biologically plausible candidate gene for female-specific genetic determination of RV function, showing associations with cardiac performance under chronically increased afterload in female patients with PAH. Further studies are needed to explore the underlying biological pathways.

## INTRODUCTION

Right and left heart structures are derived from different embryogenic cell populations and function under different physiological conditions.^1^ The right ventricle (RV) is coupled to the systemic venous return and pulmonary vascular bed, coping with relatively low pre- and afterload conditions.^1, 2^ Sex differences are common in cardiovascular physiology and diseases, and studies in populations with a low prevalence of cardiac diseases have revealed that RV volumes are smaller in women, while ejection fraction is higher compared to men.^3–6^ In conditions primarily affecting the RV, such as pulmonary arterial hypertension (PAH), where RV failure continues to be the primary cause of mortality, sex differences in RV function and adaption may explain the survival benefit of female patients.^7–9^ Unravelling the molecular mechanisms underlying sex differences in RV structure and function could directly inform novel strategies to improve RV functionality and adaptation in patients with chronically increased loading conditions. Preclinical and clinical data have linked sex hormones to the difference in RV function, and the analysis of transcriptional profiles from RV tissue has observed distinct pathways associated with RV failure in women and men.^10–15^ However, identifying these molecular mechanisms remain challenging, partly due to limited accessibility to study the human RV directly and small sample sizes.

Advances in machine learning have enabled the derivation of image-derived complex phenotypes in a manner that is scalable. This has permitted biobank-wide investigations into RV phenotypes and has revealed substantial heritability.^4–6^ We hypothesised that, considering the moderate to high heritability, the observed sex differences in RV phenotypes may be related, in part, to genetic factors. In this report, we confirmed observational evidence indicating sex differences in RV structure and function. These findings remained consistent irrespective of left heart and lung function. Through sex-stratified genetic association testing, we identified sex-specific genetic loci in *BMPR1A* (bone morphogenetic protein receptor type 1A) and near to *DMRT2* (doublesex and mab-3 related transcription factor 2), where associations with RV phenotypes were exclusively found in women. We prioritized the *BMPR1A* locus based on epigenetic data in RV tissue and validated its impact on cardiac performance in female patients with PAH.

## METHODS

### Cohort and imaging

This research has been conducted using the UK Biobank resource, a population-based cohort that recruited individuals aged 40 – 69 years in the UK from 2006 to 2010. We analysed all participants free from a history of myocardial infarction or heart failure with available genetic and imaging data, who had not withdrawn consent as of June 2023.

Cardiac magnetic resonance (CMR) imaging was performed between 2014 and 2020 across three different assessment centres. Imaging was conducted on a clinical wide bore 1.5 Tesla scanner (MAGNETOM Aera, Syngo Platform VD13A, Siemens Healthcare, Erlangen, Germany). Three long-axis cine images and a complete stack of contiguous short-axis balanced steady state free precession cine images were captured with full biventricular coverage.^16^ Morphology of the ventricles across a cardiac cycle were captured from segmented images based on an extensively validated deep-learning approach.^4, 17^ Imaging phenotypes included end-diastolic volume (EDV in ml), end-systolic volume (ESV in ml), stroke volume (SV in ml) and ejection fraction (EF in %). SV and EF were calculated from EDV and ESV. We included individuals for whom all RV measurements were available.

The study protocol was approved by the Northwest Research Ethics Committee under reference number 11/NW/0382. All participants were informed and gave written informed consent. Access was provided under application number 95677.

### Observational association

To characterize the observational relationship between RV phenotypes and sex, generalised linear models were used in R (v4.0.3). Analyses were performed on absolutes volumes and on volumes indexed for the body surface area.^18^ RV phenotypes were the dependent variable and confounders were included as covariates. The ‘basic model’ included age, standing height, weight, waist circumference and assessment centre as covariates. In sensitivity analyses, we further adjusted for left ventricular phenotypes (‘Left heart model’) and spirometry measures (‘Lung function model’). Participants with missing covariates were excluded.

*Genotyping, imputation and quality control*.

Genome-wide genotyping on UK Biobank participants was performed using two purpose-designed arrays (UK BiLEVE Axiom array and UK Biobank Axiom array). Imputation was performed using the Haplotype Reference Consortium and UK10K + 1000 Genomes reference panels.^19^ Quality control measures were performed separately in women and men. We included individuals with matched self-reported and genetic sex, without evidence of sex chromosome aneuploidy, without excessive third-degree relatives, with a sample missing call rate <0.1, with a heterozygosity to missing rate within two standard deviations and with European ethnicity. We then filtered out genotyped variants with genotyping call rate <0.95 and minor allele frequency (MAF) <0.01, and imputed variants with an INFO score <0.3 and Hardy-Weinberg equilibrium exact test P<1×10^-50^. In total, 643,281 genotyped and 9,117,541 imputed variants were included. Sex difference in allele frequencies were small (interquartile range 0.0005-0.0026)

### Heritability and genome-wide association studies

We conducted variance components analyses to assess heritability and genotypic correlation of RV traits using BOLT-REML (v2.4.1).^20^ Analyses were performed separately in women, men and the sex-combined data including hard-called genotypes (all autosomes and the X-chromosome) that passed quality control in both women and men. To assess the contribution of single chromosome to heritability, we partitioned across the chromosomes in the sex-combined dataset and repeated the analyses for each chromosome separately. Sex differences between measured heritability was assessed using Z-score and its associated P-value statistic (P_Heri_) as suggested previously.^21^

To identify single genetic variation associated with RV traits, we performed genome-wide association studies (GWAS) using BOLT-LMM (v2.4.1) which accounts for cryptic population structure and sample relatedness by fitting a linear mixed model with a Bayesian mixture prior.^22^ We used again the full panel of hard-called genotypes to construct the genetic relationship matrix. In this analysis mode, BOLT-LMM treats individuals with one X-chromosome as having an allelic dosage of 0/2 and those with two X-chromosomes as having an allelic dosage of 0/1/2. Inverse normal transformation followed by Z-scoring was applied to the RV traits before conducting GWAS. Models were adjusted for age at recruitment, age at assessment, standing height, weight, genotype array, genotype batch, imaging centre and the first 10 principal components. Principal components were generated for each sex separately using PLINK (v2.0).^23^ Variants at a commonly used significance threshold of P<5×10^−8^ were considered genome-wide significant. A genomic locus was defined as 5 Mb upstream and downstream of the variant with the lowest P-value. In each genomic locus, clumping for linkage disequilibrium (LD) was performed with an r^2^ threshold of 0.001 in the sex-combined genotypes rather than a generic reference panel.

To assess heterogeneity of GWAS loci between sexes, a fixed-effect meta-analysis framework was employed by weighting the inverse of the variance. This was performed using the ‘-sex’ option in GWAMA (v2.2.2), which provides metrics including Cochran’s Q-statistics, I^2^-statistics and P-values for assessing sex-associated heterogeneity (P_Het_).^24^ The effects of heterogeneous loci were categorized as sex-differential (effect in both sexes, but of different magnitude) or sex-specific (effect in one sex, but not the other), as suggested previously.^21^ To account for possible confounding through LD, we performed statistical colocalization and tested the hypothesis of shared genetic signal (H4) using default prior probabilities as implemented in the R package coloc (v5.1.0).^25^

### Functional annotation

Lead variants and their LD proxies (r^2^ > 0.9) were evaluated for functional consequences using the Ensembl Variant Effect Predictor tool and were assessed against publicly available epigenomic data derived from RV tissue in adult women, obtained from the ENCODE library (ENCSR821DYI, ENCSR356RNZ, ENCSR494QLL, ENCSR377RPJ) and NCBI GEO repository (GSM2322554).^26–28^ To perform epigenetic mapping, we utilized topologically associated domains from chromatin contact maps, chromatin accessibility by DNase I hypersensitive sites and histone H3 modifications. Additionally, we investigated whether variants were associated with gene expression, employing cis-expression quantitative trait loci (QTL) in cardiac tissue from the Genotype-Tissue Expression project (GTEx v7), and performed look-ups and statistical colocalization for associations with common traits, using both GWAS catalogue and PhenoScanner (v2) databases.^29–31^

### Association with the electrocardiographic PR interval

We evaluated candidate loci against electrocardiographic PR intervals separately for women and men, using the same study sample from the UK Biobank. All participants with available 12-lead electrocardiographic data were included. Generalized linear regression models in R were utilized, adjusting for age at assessment, standing height, weight, heart rate, electrocardiogram device, assessment centre, genotype array, genotype batch, and the first 10 principal components, as reported previously.^32^

### Association with cardiac index in pulmonary arterial hypertension

We validated candidate loci by assessing invasively measured cardiac performance in patients with idiopathic or heritable PAH, recruited to the UK National Institute for Health Research BioResource Rare Diseases study. All enrolled patients provided written informed consent from their respective institutions. Phenotyping, genotyping, and quality control procedures were previously reported.^33^ We included unrelated patients of European descent with available cardiac index measurements (i.e., cardiac output normalised for the body surface area, l/min/m²). Additionally, two measures of RV afterload were analysed: mean pulmonary arterial pressure (mmHg) and pulmonary vascular resistance (Wood units). Phenotypic outliers were excluded. Generalised linear regression was employed to examine single-marker variants for genetic associations with hemodynamic measures using R. Analyses were adjusted for age at diagnosis, read length chemistry, and the first 10 principal components. Principal components were again generated for each sex separately using PLINK.

## RESULTS

We investigated 18,156 women and 16,171 men of European ancestry free from pre-existing heart disease (combined 34,327 individuals). Most women (93.5%) were post-menopausal. Main characteristics of the cohort are reported in supplementary table S1.

### Observational associations between RV phenotypes and sex

The absolute differences in means between women and men were -47.4 ml (95% confidence interval [CI] -48.4 - -46.8) for RVEDV, -26.7 ml (95% CI -27.1 - -26.4) for RVESV, -20.6 ml (95% CI -21 - -20.3) for RVSV and 4.1 % (95% CI 4 - 4.2) for RVEF (Figure S1). Differences were similar for RV volumes indexed to the body surface area with -13.8 ml/m² (95% CI -14.1 - -13.5) for RVEDVI, -9.3 ml/m² (95% CI -9.5 - -9.1) for RVESVI and -4.5 ml/m² (95% CI -4.7 - -4.3) for RVSVI (Figure S2). Sex was independently associated with RV phenotypes in multivariable regression models accounting for left ventricular counterparts and pulmonary function. Multivariate models were adjusted for height and weight, and indexing the phenotypes to body surface area did not change results (Table 1).

**Table 1:** Effect estimates of female sex on right ventricular (RV) imaging traits in multivariable linear regression models.

| Phenotype | Unit* | Basic model<sup>†</sup> |  |  | Left heart model<sup>‡</sup> |  |  | Lung function model<sup>§</sup> |  |  |
| --- | --- | --- | --- | --- | --- | --- | --- | --- | --- | --- |
| | | $\beta$ | 95% CI | P | $\beta$ | 95% CI | P | $\beta$ | 95% CI | P |
| RVEDV | ml | -27.8 | -28.6 - -26.9 | <0.001 | -11.5 | -12 - -10.9 | <0.001 | -24.8 | -25.7 - -23.8 | <0.001 |
|  | ml/m<sup>2</sup> | -14.5 | -14.9 - -14.1 | <0.001 | -6.1 | -6.4 - -5.8 | <0.001 | -13.1 | -13.5 - -12.6 | <0.001 |
| RVESV | ml | -17.3 | -17.8 - -16.8 | <0.001 | -9.4 | -9.8 - -9 | <0.001 | -16.1 | -16.7 - -15.6 | <0.001 |
|  | ml/m<sup>2</sup> | -9.1 | -9.3 - -8.8 | <0.001 | -4.9 | -5.1 - -4.7 | <0.001 | -8.5 | -8.8 - -8.2 | <0.001 |
| RVSV | ml | -10.5 | -11 - -9.9 | <0.001 | -4.2 | -4.5 - -3.8 | <0.001 | -8.6 | -9.2 - -8 | <0.001 |
|  | ml/m<sup>2</sup> | -5.4 | -5.7 - -5.2 | <0.001 | -2.2 | -2.4 - -2 | <0.001 | -4.5 | -4.8 - -4.2 | <0.001 |
| RVEF | % | 3.4 | 3.2 – 3.6 | <0.001 | 2 | 1.8 – 2.2 | <0.001 | 3.5 | 3.3 – 3.8 | <0.001 |

\* RV volumes were assessed as either absolute measures in ml or indexed for the body surface area in ml/m<sup>2</sup>.
<sup>†</sup> Basic model includes age, standing height, weight, waist circumference and assessment centre.
<sup>‡</sup> Left heart model includes the basic model and the LV counterpart (LVEDV, LVESV, LVSV or LVEF; either as absolute value of or indexed for the body surface area).
<sup>§</sup> Lung function model includes the basic model and spirometry (FEV1, FVC and FEV1 to FVC ratio).
Confidence interval (CI). Forced expiratory volume in 1 second (FEV1). Forced vital capacity (FVC). Left ventricle (LV). LV end-diastolic volume (LVEDV). LV end-systolic volume (LVESV). LV stroke volume (LVSV). LV ejection fraction (LVEF). RV end-diastolic volume (RVEDV). RV end-systolic volume (RVESV). RV stroke volume (RVSV). RV ejection fraction (RVEF).

Models adjusted for left ventricular phenotypes revealed the smallest effect sizes, suggesting that ventricular interdependence might partially mediate the associations. We therefore tested whether sex differences in left heart phenotypes account for the RV differences. Three RV phenotypes (RVEDV, RVESV and RVEF) remained statistically significant associated with sex after introducing the interaction term between left ventricular counterpart and sex into the model (Table S2). This suggests that while RVEDV and RVESV were smaller in women, a similar RVSV between the sexes resulted in a better RVEF in the female population.

### Comparisons of heritability and genotypic correlations between sexes

Structural phenotypes of RV showed substantial heritability, which was as high as 43% in women and 44% in men for RVEDV. The functional phenotype, RVEF, displayed lower heritability in both sexes (24%). Overall, there were no differences in the measured heritability between sexes (P_Heri_>0.05; Figure S3). Furthermore, no individual chromosome, including the X-chromosome, contributed to the overall heritability more or less than expected given its length and number of variants (Figure S4).

Genotypic correlations were strong between RVEDV and RVSV in both women and men (r_g_=0.83). The strongest negative correlations were observed between RVESV and RVEF, with r_g_ of -0.72 in women and -0.66 in men. Overall, the pattern of genotypic correlations between RV phenotypes was comparably in both sexes (Table S3).

### Identification of individual genotypic effects

We conducted GWAS on RV phenotypes in women, men and both sexes combined. In total, we identified 68 putative susceptibility loci at a commonly used significance threshold of P<5×10^−8^ after clumping to remove variants in LD (r^2^>0.001). These included 10 loci in women, 12 in men and 46 in the sex-combined analysis. Manhattan plots for all GWAS are depicted in the supplement (Figures S5 to S8). No loci were found on the X-chromosome. Multiple loci were common to both sexes, resulting in 48 unique pairs of associations between RV phenotypes and genomic regions. Similarly, several loci were shared among different RV phenotypes, and 48 unique association pairs mapped onto 28 distinct genomic regions following LD clumping across phenotypes (Table S4). In previous investigations, GWAS of RV phenotypes combining data from both sexes were conducted on the same set of CMR images, employing distinct analytical pipelines.^5, 6^ All 48 unique pairs between RV phenotypes and genomic regions identified in the present analyses showed consistent allelic effect directions and achieved statistical significance (P<0.05) in at least one of the previous studies, indicating robust technical reproducibility (Table S5).

### Identification of heterogeneity of allelic effects between sexes

We next tested for heterogeneity of allelic effects between women and men. Among 48 unique association pairs, six pairs (13%) revealed significant heterogeneity between sexes (P_Het_<0.05, Figure 1A-C). These six pairs mapped to five distinct genomic regions with nearest protein-coding genes being *LSM3* (homolog, U6 small nuclear RNA and mRNA degradation associated), *RSRC1* (arginine and serine rich coiled-coil 1), *NOS3* (nitric oxide synthase 3), *DMRT2* and *BMPR1A* (Table 2). In four association pairs, genetic effects in the sex-combined analyses were driven by one sex due to unequal effect sizes between women and men (referred to as sex-differential; Figure 1D). Stronger signals for sex-differential effects were observed in men for RVEF near *LSM3* and for RVSV and RVEDV near *NOS3*. The association between RVEF and variants in *RSRC1* showed a stronger effect in women compared to men (Table 2). At the *RSRC1* locus, lead variants with the most significant p-values were different between sexes suggesting different functional variants (posterior probability for a shared causal variant was only 1.7%; Figure S9).

**Figure 1:**
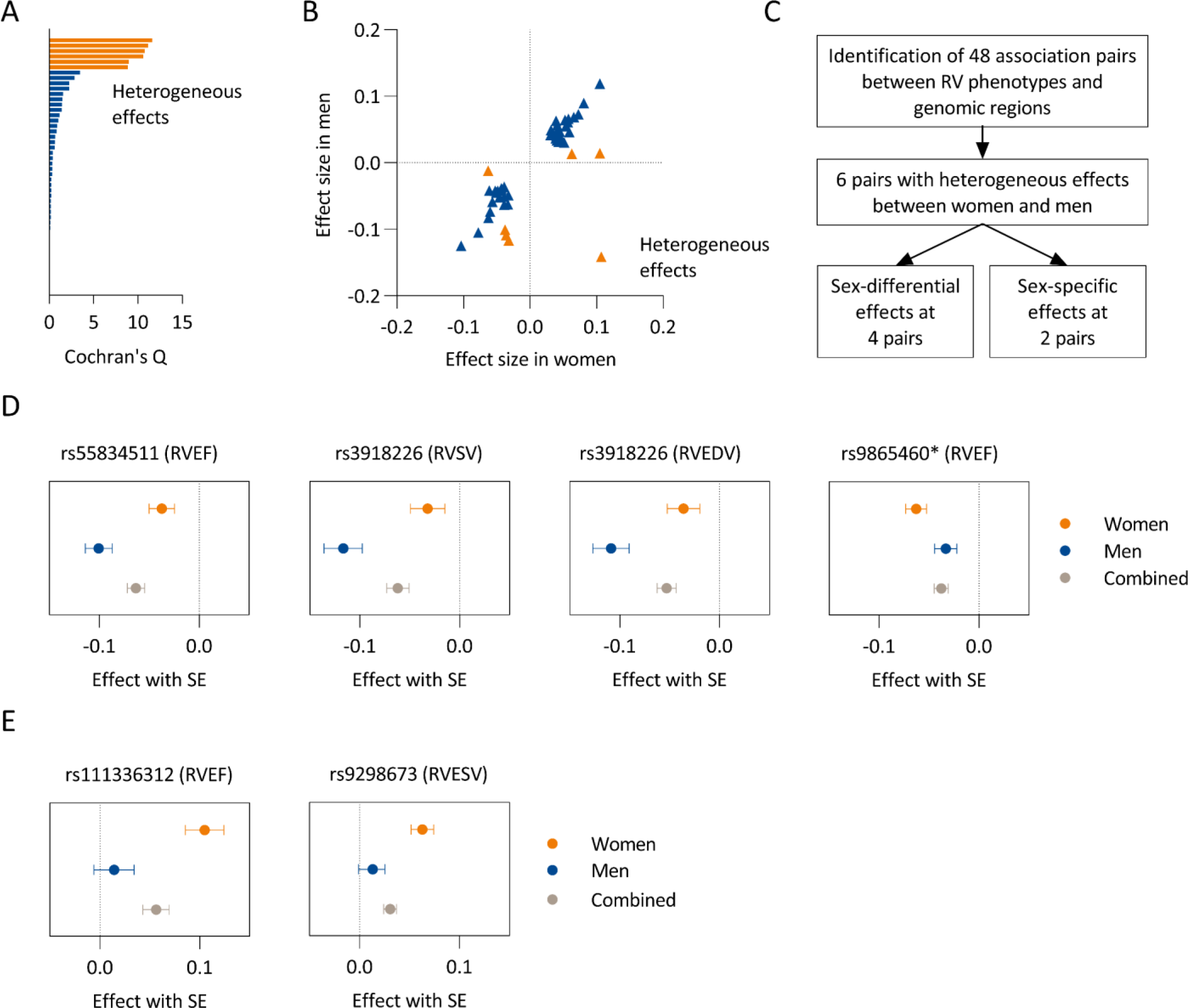
Heterogeneity in allelic effects between women and men. In **A** and **B**, six association pairs were highlighted that showed significant heterogeneity between women and men. The effects of these six association pairs were distinguished into four sex-differential effects, where the combined effect was predominantly driven by one sex, and two sex-specific effects, which were present in only one sex (**C**). Sex-differential allelic effects are depicted in **D** and sex-specific effects in **E**. The asterisk indicate that in men rs3851363 (r^2^ 0.36 with lead variant in women) was used due to signal differences between women and men. Right ventricle (RV). RV end-diastolic volume (RVEDV). RV end-systolic volume (RVESV). RV stroke volume (RVSV). RV ejection fraction (RVEF). Standard error (SE).

**Table 2:**
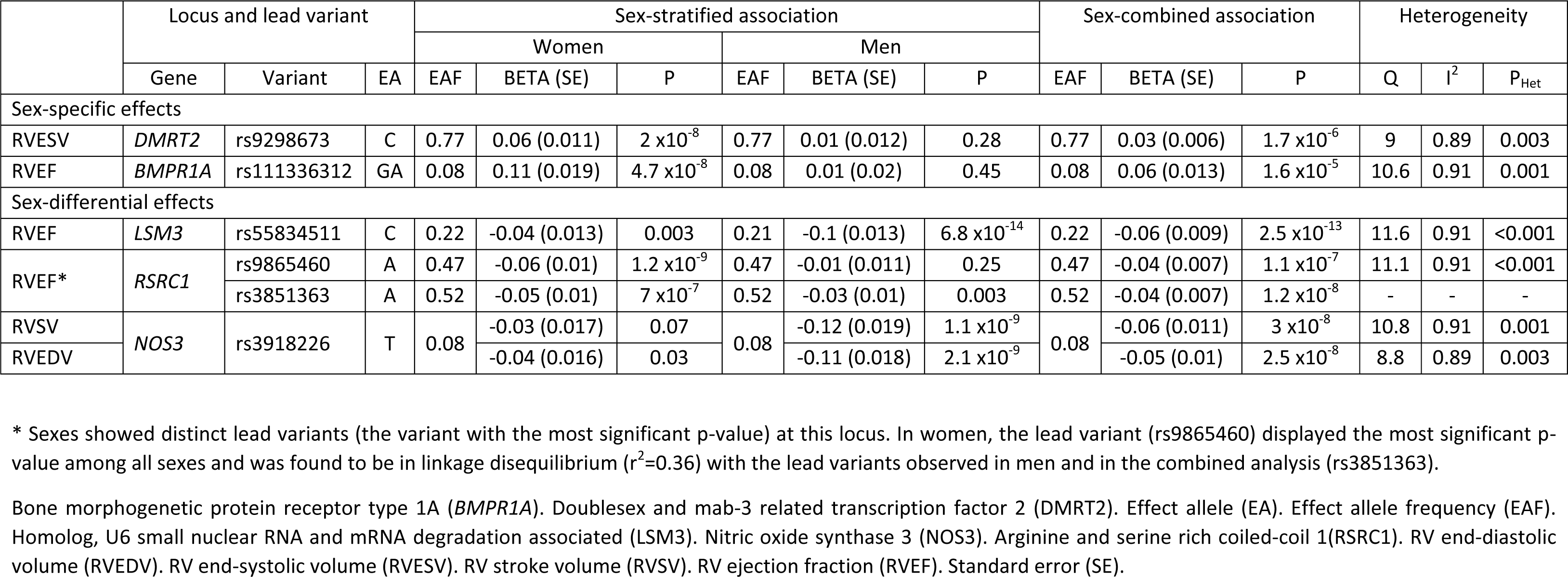
Genetic loci with heterogeneous effects between women and men on right ventricular (RV) phenotypes.

We identified two association pairs that were only present in women (referred to as sex-specific; Figure 1E). Our sex-combined analyses failed to detect genome-wide significant signals at these loci (Table 3). Likewise, these loci only reached nominal significance in previous data combining sexes (Table S4). Specifically, we observed sex-specific effects in women for variants located in *BMPR1A* on RVEF (lead variant rs111336312) and for a genomic region upstream of *DMRT2* on RVESV (lead variant rs9298673). No association signals were found at these two regions in men (Figure 2A-B).

**Figure 2:**
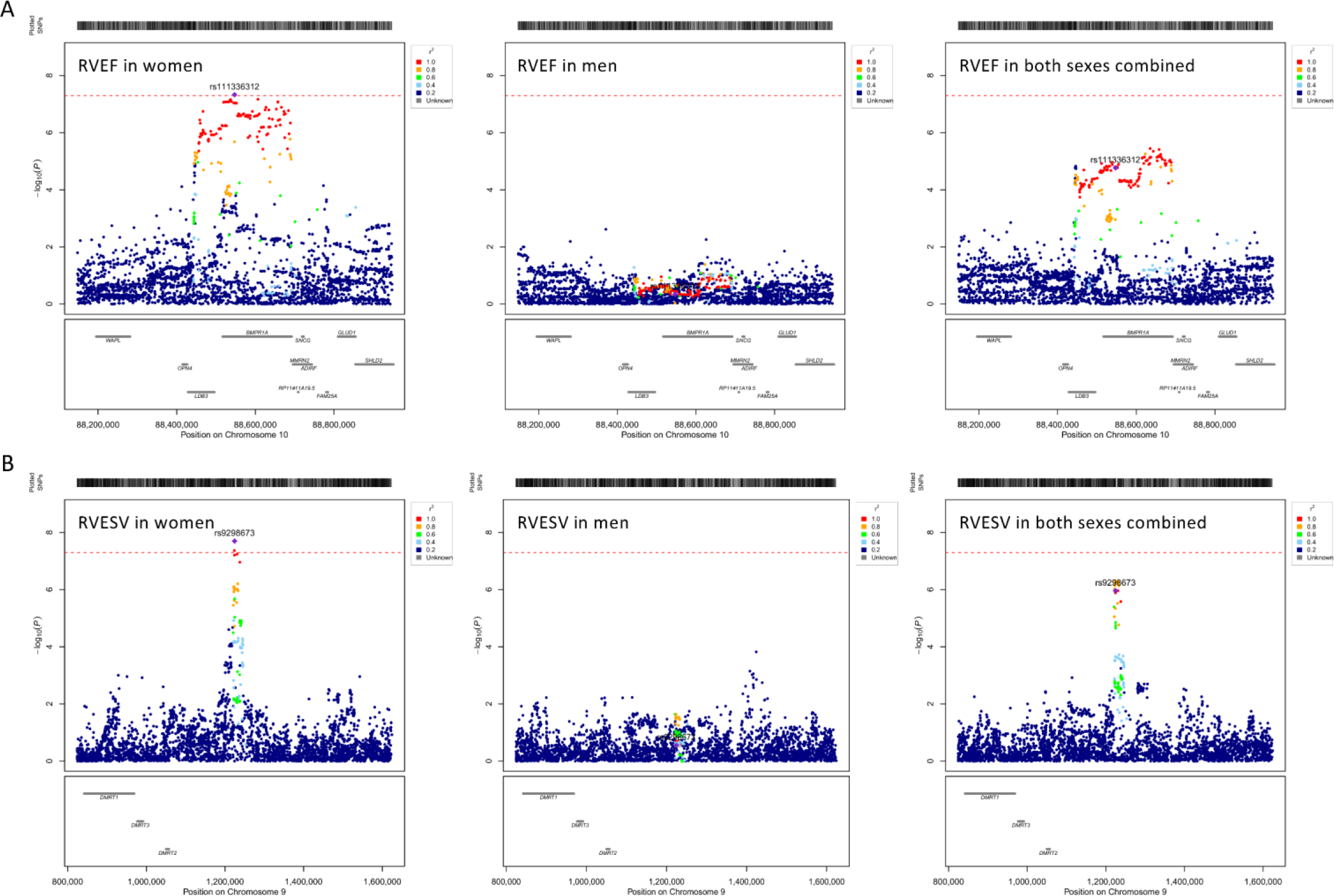
Regional association for sex-specific loci. Regional plots show the genomic positions of variants on the x-axis and their association strength (-log10(p-value)) on the y-axis. Correlation coefficients (r²) with the lead variants are indicated by colour. In **A**, the locus for RVEF on chromosome 10 is displayed. In **B**, the locus for RVESV on chromosome 9. RV end-systolic volume (RVESV). RV ejection fraction (RVEF).

**Table 3:**
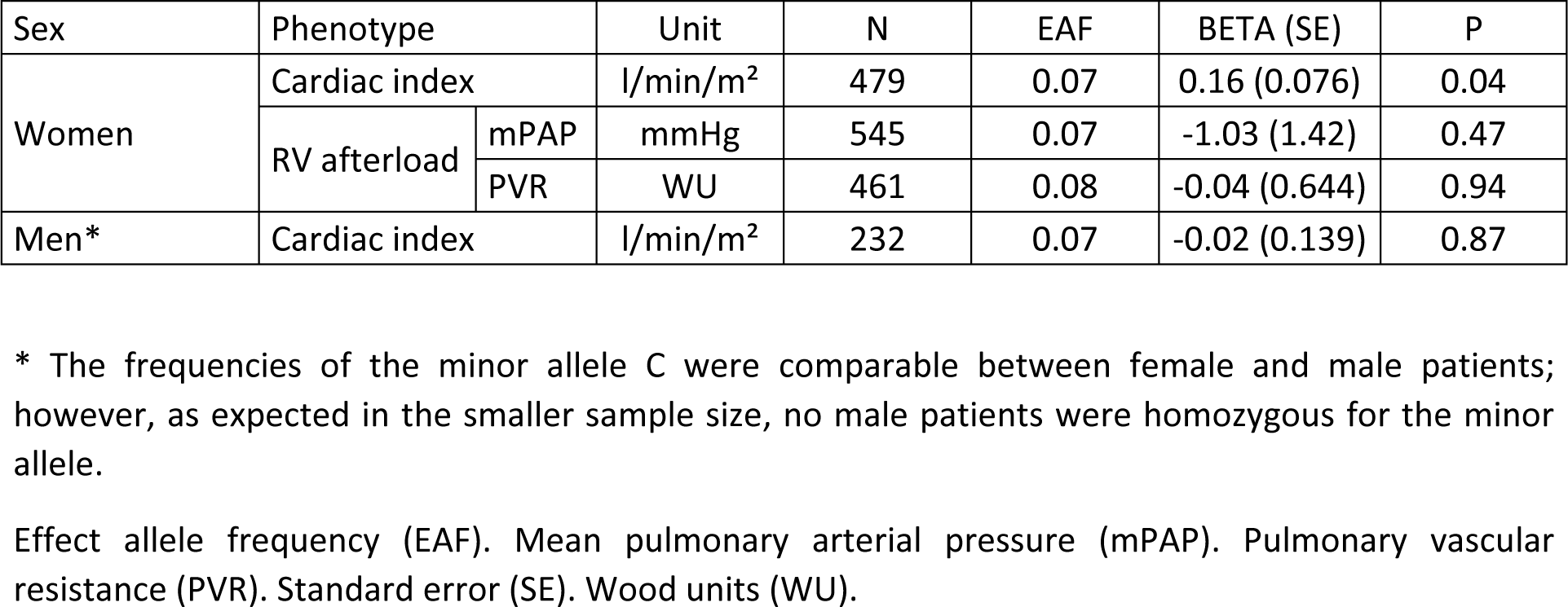
Genotypic effect of each minor allele C at rs140745739 on hemodynamic measures in patients with idiopathic or hereditable pulmonary arterial hypertension (PAH).

### Functional annotation of sex-specific loci

The RV phenotype-associated variants at the sex-specific loci in *BMPR1A* and near *DMRT2* were not protein-coding. We therefore searched for evidence of regulatory elements in publicly available epigenomic data from RV tissue obtained from three different adult women. Variants associated with RVEF in women extend across the introns of *BMPR1A* and its promoter. Among these variants, rs140745739, which is highly linked to the lead variant (r²=0.97), resides directly within the promoter region, marked by a DNase I hypersensitivity indicating accessibility to the binding of transcription factors. Histone H3 lysine 4 trimethylation and lysine 27 acetylation modifications flank the promotor region, indicating active transcription (Figure 3A). In line with this, the *BMPR1A* locus (including rs140745739) has previously been associated with significant cis-expression QTLs for *BMPR1A* in cardiac tissue from both sexes.^29^

**Figure 3:**
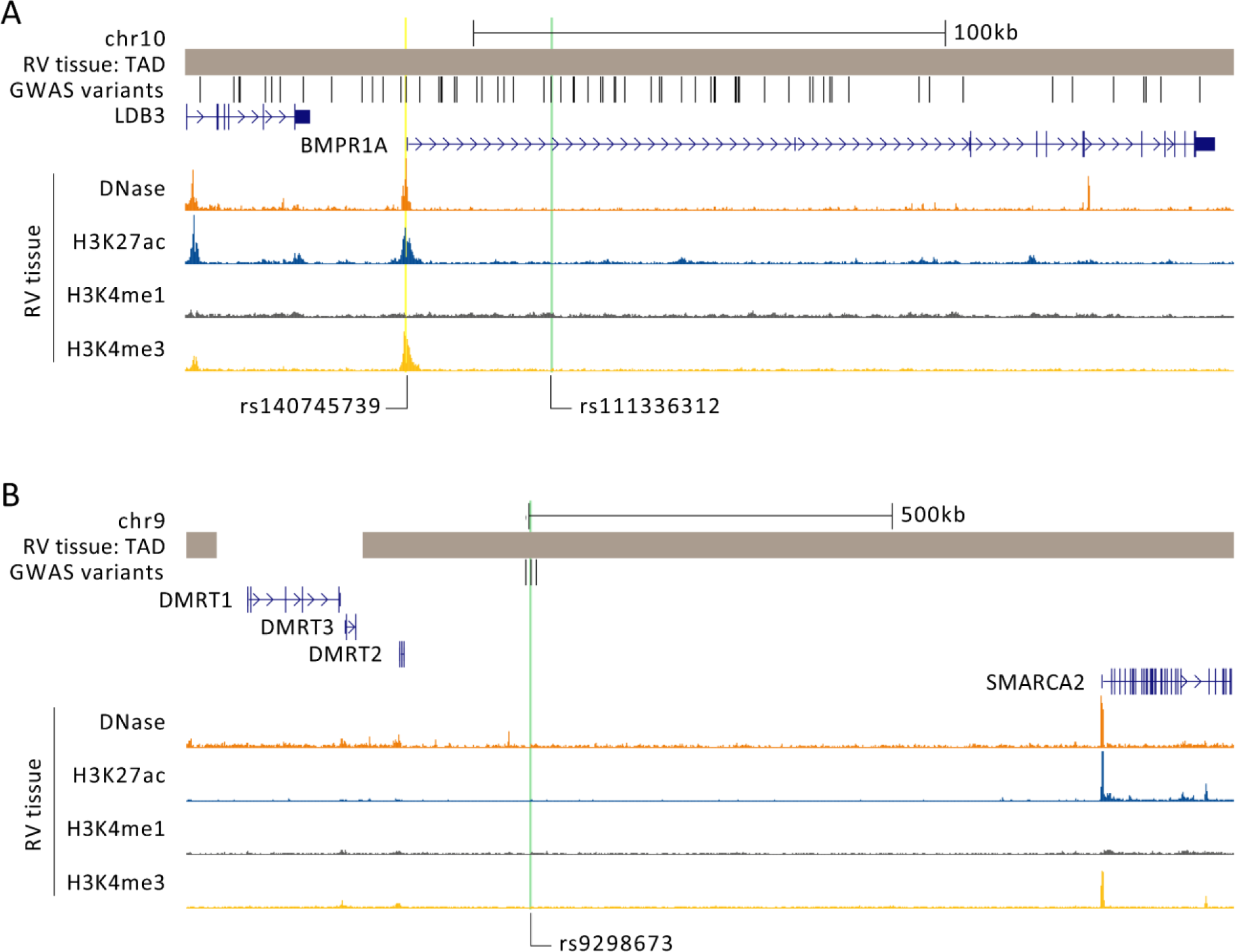
Epigenetic marks in right ventricular (RV) tissue from adult women. Topologically associated domains (TAD) indicate genomic regions that are in close proximity based on the 3-dimensional chromatin structure (Hi-C maps). GWAS variants are displayed that are in high LD with the lead variant (r^2^>0.9). Lead variants are highlighted in green. DNase I hypersenstivity sites mark chromatin regions that are assessable for transcription factors. Epigenomic modification indicate areas likely to contain active regulatory regions and promoters. H3K4Me1 is often found in enhancers, H3K4Me3 typically marks promoters and H3K27ac is often marking active regulatory regions. In **A**, the female-specific locus for RVEF on chromosome 10 is displayed. Here a variant rs140745739 (highlighted in yellow) that is in high LD with lead variant (r²=0.97) resides within the active promotor region of BMPR1A. In **B**, the female-specific locus for RVESV on chromosome 9 is displayed. DMRT2 is the closest gene and shares a TAD with the GWAS variants but no adjacent active regulatory region is found. Bone morphogenetic protein receptor type 1A (BMPR1A). Doublesex and mab3-related transcription factor 1 to 3 (DMRTs). Histone H3 lysine 4 monomethylation (H3K4Me1). Histone H3 lysine 4 trimethylation (H3K4Me3). H3 lysine 27 acetylation (H3K27ac). LIM Domain Binding 3 (LDB3). RV end-systolic volume (RVESV). RV ejection fraction (RVEF). SWI/SNF Related, Matrix Associated, Actin Dependent Regulator Of Chromatin, Subfamily A, Member 2 (SMARCA2).

The locus that associated with RVESV in women is located within the intergenic space between *DMRT2* (the nearest gene) and *SMARCA2* (SWI/SNF related, matrix associated, actin dependent regulator of chromatin, subfamily A, member 2) and shares a topologically associated domain with both genes. Based on the available data, we did not identify evidence of an active regulatory element in RV tissue within the region containing the GWAS variants (Figure 3B).

### Effect of BMPR1A locus on electrocardiographic PR interval

Annotation to common traits revealed that the *BMPR1A* locus identified in our study colocalised with a locus for the electrocardiographic PR interval (posterior probability for a shared causal variant was 86%; Figure S10).^32^ We subsequently assessed the association of rs140745739, located within the *BMPR1A* promoter, with the PR interval in our study cohort. The PR interval was available for 15,287 women and 14,002 men. The minor allele C was significantly associated with a shorter PR interval in women (effect size -1.62, standard error 0.512, P=0.002), while no association was observed in men (effect size -0.4, standard error 0.622, P=0.53), emphasizing further a female-specific cardiac phenotype.

### Effect of BMPR1A locus on cardiac performance in PAH

PAH is characterised by chronically increased RV afterload that can compromise RVEF, followed by a reduction in cardiac index. To assess the effect of genetic variation at the *BMPR1A* locus on cardiac performance in patients with idiopathic or hereditable PAH, we correlated rs140745739 genotype with cardiac index. In 479 female patients with PAH, the minor allele C was significantly associated with an elevated cardiac index (effect size 0.16), aligning with its association with increased RVEF in UK Biobank (Figure 4A). We did not detect an association with mean pulmonary arterial pressure or pulmonary vascular resistance, indicating that the elevation in cardiac index was likely not explained by altered RV afterload. Additionally, we did not detect an effect on cardiac index in 232 male patients with PAH (effect size -0.02), despite a similar allele frequency (Table 3).

**Figure 4:**
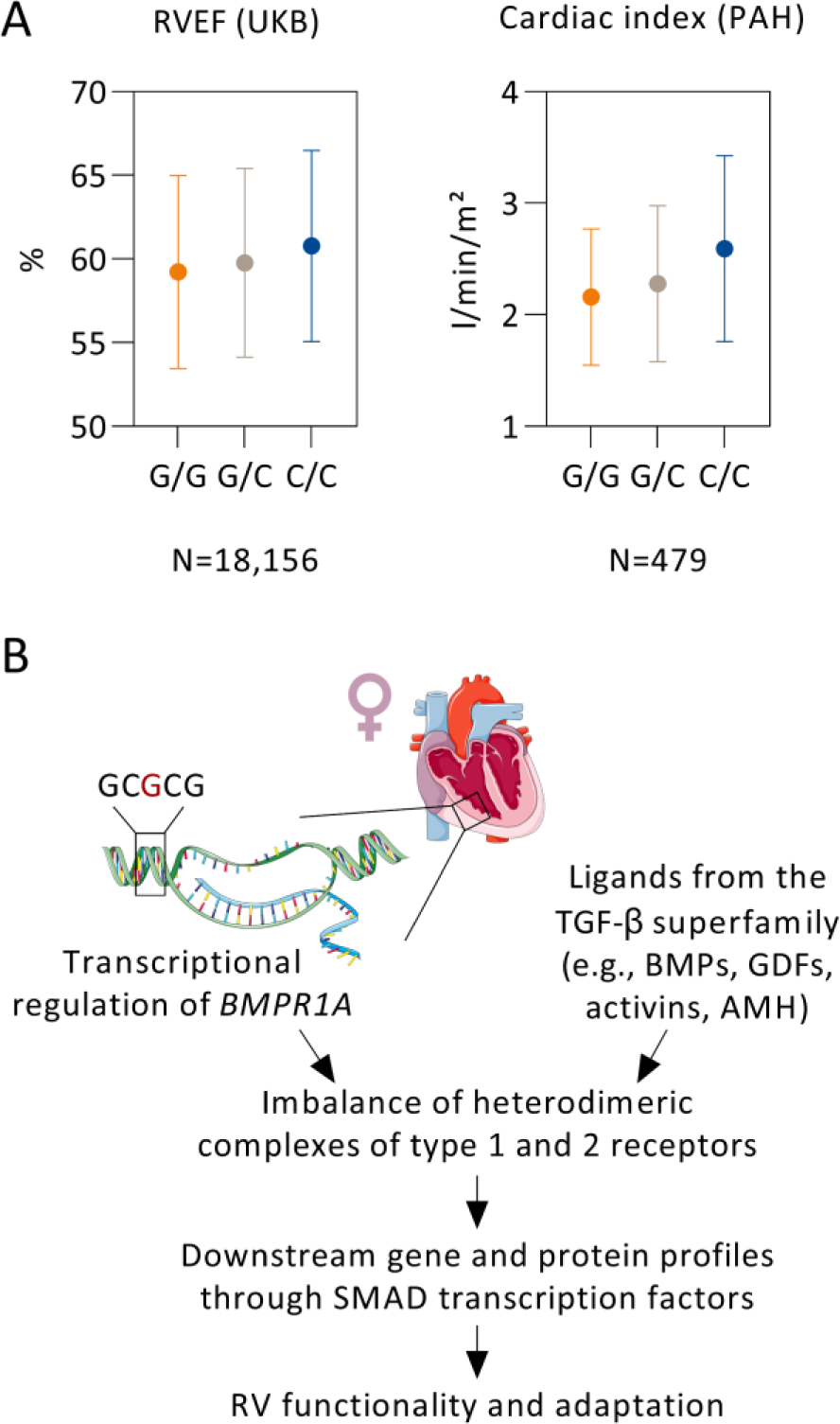
*BMPR1A* locus in women. In **A**, aligned genotypic effects of the BMPR1A promoter variant (rs140745739, minor allele C) on right ventricular ejection fraction (RVEF) in 18,156 female participants of the UK Biobank (UKB) and on cardiac index in 479 female patients with idiopathic or hereditary pulmonary arterial hypertension (PAH). Data are presented as the mean with standard deviation. In **B**, a schematic depicts potential mechanisms of BMPR1A in RV biology in women. Anti-Müllerian hormone (AMH). Bone morphogenetic proteins (BMPs). Bone morphogenetic protein receptor type 1A (BMPR1A). Growth differentiation factors (GDFs). Transforming growth factor (TGF).

## DISCUSSION

We examined sex-stratified genetic determinants of clinically relevant RV structural and functional phenotypes derived from 34,327 CMR examinations in UK Biobank. We confirmed that women had smaller RV volumes and higher ejection fraction, which were independent of left ventricular counterparts and pulmonary function. Two sex-specific genetic associations were identified. In women, common genetic variants near the promoter of *BMPR1A*, likely affecting *BMPR1A* transcription, were identified as candidate genetic determinants of RVEF. Variants located upstream of *DMRT2* were identified as candidates influencing RVESV in women. Further investigation of female patients with PAH shows that the minor C allele at rs140745739, located in the *BMPR1A* promoter, is associated with a better cardiac index despite similar RV afterload, consistent with its association with increased RVEF in the UK Biobank cohort.

*BMPR1A* (also known as ALK3) is a biologically plausible candidate for a role in RV biology (Figure 4B). In the developing heart, *BMPR1A* is required for the expansion of the early second heart field, which subsequently contributes to growth of the RV.^34^ During later embryogenesis, selective deletion of *BMPR1A* in cardiac myocytes results in endocardial cushion defects and myocardial thinning.^35^ In adult mice, incomplete loss of *BMPR1A* in cardiac myocytes results in sporadically depressed RV myocardial function.^36^ In line with these observations, rare loss-of-function mutations in *BMPR1A* have been identified in patients with congenital heart diseases.^37^ Furthermore, BMPR1A signalling is required for normal AV node conduction, supporting a functional role of *BMPR1A* as a genetic determinant of the PR interval.^32, 38^ The *BMPR1A* locus associated with RVEF in women and PR prolongation spans across the introns of *BMPR1A* and its promoter region. Epigenetic marks suggest active transcription of *BMPR1A* in RV tissue, and the location of genetic variants in the promoter region indicates a regulatory function. This interpretation is supported by the association of these variants with the *BMPR1A* transcript level in cardiac tissue.^29^ The *BMPR1A* variants may be acting on PR interval and RVEF independently but there may also be a physiological link; longer PR intervals can have negative hemodynamic effects as a result of diastolic atrioventricular valve regurgitation, decreased ventricular preload, and a smaller SV.^39–41^

On a cellular level, BMPR1A functions as a type I receptor in the BMP signalling pathway, where it combines with the type II receptor BMPR2 (bone morphogenetic protein receptor type 2) to bind ligands that signal through SMAD transcriptional regulators. Regulation of *BMPR1A* expression has been linked to candidate transcription factors through in silico analysis, and the rs140745739 variant is located within binding signals from ChIP-seq experiments in various cell types.^28, 42^ The expression of *BMPR1A* is reduced in the failing RV from PAH patients, suggesting that a therapeutic strategy to increase BMPR1A expression would be beneficial.^43^ Preclinical data indicate that BMP7-based small peptides, acting as activators of BMPR1A, might offer protection against pathological cardiac remodelling induced by pressure overload.^44^ Additionally, BMPR1A has been identified as a receptor for AMH (anti-Müllerian hormone), produced by ovarian follicles in the early stages of cyclic development.^45^ AMH has been detected in peripheral circulation, and higher levels have been linked with cardiovascular health in adult women.^46^ Understanding cardiac BMP signalling in women could provide novel therapeutic strategies aimed at improving RV function in a group of patient that most likely to benefit from therapeutic interference.

The second sex-specific signal in women is situated in the intergenic region, between *DMRT2* and *SMARCA2*. Both genes have been implicated in organ development, but their specific roles in the cardiovascular system remain unclear. The absence of epigenetic marks in RV tissue associated with the GWAS loci emerging in our data may indeed suggest a regulatory function in non-cardiac cell systems. The *DMRT* family of genes are highly conserved transcription factors that play roles in sexual regulation, including sex differentiation, sexual dimorphism, and spermatogenesis. *DMRT* genes are also involved in other developmental processes such as myogenesis. Notably, common genetic variants near *DMRT2* (not in LD with our GWAS variants) have been linked to skeletal muscle traits in cardiorespiratory exercise testing.^47^ *SMARCA2* is a member of the ATP-dependent SWI/SNF complex, which modulates chromatin structure to facilitate gene transcription. Within the cardiovascular system, *SMARCA2*-mediated catalysis of the SWI/SNF complexes is required for maintaining the differentiation of vascular endothelial cells in adults.^48^

Several potential mechanisms may underlie genetic determinants of phenotypic differences between women and men. These mechanisms encompass the influence of sex hormones with downstream transcriptional regulation, sex-related QTL affecting both gene expression and protein expression, variations in gene splicing, and epigenetic control. Sex differences in gene expression across different tissues has been identified in around 37% of human genes.^49^ In contrast, sex-biased cis-QTL for gene expression were less frequent but were able to inform GWAS hits, suggesting that sex-specific genetic effects on gene expression are present for a subset of previously identified cis-expression QTLs, and some of these sex-biased QTL influence genes linked to human phenotypes.^49^

We acknowledge limitations in our study. Our study focused on individuals of European descent and generalizability of the findings to non-European populations remains uncertain. There is overrepresentation of female participants from the UK biobank (with a ratio of 1.3:1), which may have contributed to the detection of genetic signals. Similarly, the PAH cohort was predominantly female.^33^ Although our study provided statistical support for sex-specific loci associated with RV phenotypes, further experimental research, such as gene-editing, is required to validate our findings and annotate genetic variants with biologically meaningful functions.

In summary, our study provides observational evidence regarding the relationship between sex and RV phenotypes and evidence for sex-differences in genetic determinants, with *BMPR1A* as a priority candidate. Performing sex-stratified genetic association analyses may contribute to our understanding of the genetic basis of phenotypic sex differences and disease outcomes. Further efforts are warranted to explore the underlying biological factors.

## Data Availability

The raw imaging data, genotype data and non-imaging participant characteristics are available from UK Biobank via a standard application procedure at http://www.ukbiobank.ac.uk/. Summary GWAS statistics will be made available upon publication. Further web links for the publicly available datasets used in the study are as follows: PhenoScanner (http://www.phenoscanner.medschl.cam.ac.uk), GTEx (https://gtexportal.org), Ensembl VEP (https://www.ensembl.org/info/docs/tools/vep), ENCODE (https://www.encodeproject.org), and NCBI GEO (https://www.ncbi.nlm.nih.gov/geo/). All other data are contained in the article file and its supplementary information or are available upon request.

## ABBREVIATIONS

AMH: Anti-Müllerian hormone
BMP: Bone morphogenetic protein
BMPR1A: Bone morphogenetic protein receptor type 1A
CMR: Cardiac magnetic resonance
DMRT2: Doublesex and mab-3 related transcription factor 2
EDV: End-diastolic volume
EF: Ejection fraction
ESV: End-systolic volume
GWAS: Genome-wide association studies
LD: Linkage disequilibrium
LSM3: Homolog, U6 small nuclear RNA and mRNA degradation associated
MAF: Minor allele frequency
NOS3: Nitric oxide synthase 3
PAH: Pulmonary arterial hypertension
QTL: Quantitative trait locus
RSRC1: Arginine and serine rich coiled-coil 1
RV: Right ventricle
SV: Stroke volume

## ACKNOWLEDGMENT

The authors thank all volunteers and patients for their participation and acknowledge the UK Biobank and NIHR BioResource centres and staff for their contributions. We thank the NIHR BioResource – Rare Diseases Consortium and UK PAH Cohort Study Consortium as reported in ^33^ for data access. In addition, we thank Professor Renate B. Schnabel and Carla Reinbold for their review of the data presentation and statistical methods (both from the University Medical Centre Hamburg-Eppendorf).

## SUPPLEMENTAL MATERIAL

This manuscript has associated supplemental tables (S1 to S5) and figures (S1 to S10).

