## Supplemental Tables and Figures for "Sex-specific genetic determinants of right ventricular structure and function"

SUPPLEMENT

**Table S1:** Characteristics of participants when attending imaging

| | | Unit | Women | Men | Combined |
|---|---|---|---|---|---|
| | | | (N=18,156) | (N=16,171) | (N=34,327) |
| Age | | years | 63 (7.4) | 67 (7.6) | 63.6 (7.5) |
| BMI | | kg/m² | 26 (4.7) | 27 (3.9) | 27 (4.3) |
| BSA | | m² | 1.76 (0.17) | 2.02 (0.18) | 1.88 (0.22) |
| FEV1 / FVC* | | % | 75 (0.9) | 74 (0.9) | 74 (0.9) |
| SBP† | | mmHg | 135 (19) | 142 (17) | 138 (18) |
| DBP† | | mmHg | 77 (10) | 81 (10) | 79 (10) |
| RV | EDV | ml | 134 (24) | 182 (33) | 157 (37) |
| | | ml/m² | 76 (12) | 90 (16) | 83 (15) |
| | ESV | ml | 55 (13) | 82 (19) | 67 (21) |
| | | ml/m² | 31 (7) | 40 (9) | 36 (9) |
| | SV | ml | 79 (15) | 100 (20) | 89 (20) |
| | | ml/m² | 45 (8) | 50 (10) | 47 (9) |
| | EF [%] | % | 59 (6) | 55 (6) | 57 (6) |
| LV | EDV | ml | 129 (22) | 168 (31) | 148 (33) |
| | | ml/m² | 73 (11) | 83 (15) | 78 (14) |
| | ESV | ml | 50 (12) | 71 (18) | 60 (19) |
| | | ml/m² | 29 (7) | 35 (9) | 31 (8) |
| | SV | ml | 79 (15) | 97 (19) | 88 (19) |
| | | ml/m² | 45 (7) | 48 (9) | 46 (8) |
| | EF [%] | % | 61 (6) | 58 (6) | 60 (6) |

Data are presented as mean (with standard deviation) or percentage.

* Spirometry measurements were available in 16,317 women and 14,282 men (combined 30,599)

† Blood pressure measurements were available in 14,051 women and 12,679 men (combined 26,730)

Body surface area (BSA). Body mass index (BMI). Diastolic blood pressure (DBP). Forced expiratory volume in 1 second (FEV1). Forced vital capacity (FVC). Right ventricle (RV). RV end-diastolic volume (RVEDV). RV end-systolic volume (RVESV). RV stroke volume (RVSV). RV ejection fraction (RVEF). Systolic blood pressure (SBP)

**Table S2:** Effect estimates of female sex on right ventricular (RV) imaging phenotypes in multivariable linear regression

| Phenotype | Unit[#] | Left heart model with interaction term[‡] | | |
|---|---|---|---|---|
| | | β | 95% CI | P |
| RVEDV | ml | -7.8 | -9.8 - -5.8 | 6.21E-14 |
| | ml/m² | -2.9 | -4 - -1.7 | 9.63E-07 |
| RVESV | ml | -7.2 | -8.3 - -6.2 | 3.49E-40 |
| | ml/m² | -3.8 | -4.4 - -3.2 | 1.65E-37 |
| RVSV | ml | -1.4 | -2.6 - -0.1 | 3.99E-02 |
| | ml/m² | 0.5 | -0.2 - 1.3 | 1.36E-01 |
| RVEF | % | 1.7 | 0.6 - 2.8 | 1.88E-03 |

[#] RV volumes were assessed as either absolute measures in ml or indexed for the body surface area in ml/m².

[‡] Left heart model with interaction term includes age at assessment, standing height, weight, waist circumference, assessment centre, left ventricular (LV) counterpart (as absolute volume of indexed for body surface area) and the interaction term between LV counterpart and sex (LV phenotypes*sex).

Confidence interval (CI). RV end-diastolic volume (RVEDV). RV end-systolic volume (RVESV). RV stroke volume (RVSV). RV ejection fraction (RVEF).

**Table S3:** Genotypic correlations between right ventricular (RV) phenotypes within women, men and sex-combined data.

| Women | | | | |
|---|---|---|---|---|
| | RVEF | RVESV | RVEDV | RVSV |
| RVEF | | -0.72 (0.032) | -0.12 (0.062) | 0.45 (0.062) |
| RVESV | | | 0.77 (0.019) | 0.29 (0.054) |
| RVEDV | | | | 0.83 (0.021) |
| Men | | | | |
| | RVEF | RVESV | RVEDV | RVSV |
| RVEF | | -0.66 (0.048) | -0.088 (0.093) | 0.49 (0.096) |
| RVESV | | | 0.81(0.024) | 0.33 (0.076) |
| RVEDV | | | | 0.86 (0.032) |
| Sex-combined analyses | | | | |
| | RVEF | RVESV | RVEDV | RVSV |
| RVEF | | -0.69 (0.021) | -0.1 (0.039) | 0.47 (0.039) |
| RVESV | | | 0.76 (0.012) | 0.31 (0.037) |
| RVEDV | | | | 0.82 (0.015) |

Presented as genetic correlation with standard error.

RV end-diastolic volume (RVEDV). RV end-diastolic volume (RVEDV). RV end-systolic volume (RVESV). RV stroke volume (RVSV). RV ejection fraction (RVEF).

**Table S4:** Association pairs between right ventricular (RV) phenotype and genetic variants identified with summary statistics and heterogeneity measures.

| Locus, phenotype, variant | | | | | | | | | Women | | | | | Men | | | | | Combined | | | | | Heterogeneity | | | |
|---|---|---|---|---|---|---|---|---|---|---|---|---|---|---|---|---|---|---|---|---|---|---|---|---|---|---|---|
| Locus | Pheno | Sex* | RSID | CHR | BP (GRCh37) | EA | OA | INFO | EAF | Beta | SE | CHISQ | P | EAF | Beta | SE | CHISQ | P | EAF | Beta | SE | CHISQ | P | Q | P (Q) | i2 | P (Het) |
| 1 | RVEF | both | rs9442214 | 1 | 16349460 | C | T | 1 | 0.67 | -0.05 | 0.011 | 17.5 | 2.80E-05 | 0.67 | -0.05 | 0.012 | 20.1 | 7.30E-06 | 0.67 | -0.05 | 0.007 | 37.7 | 8.40E-10 | 0.1 | 0.6998 | 0 | 0.6998 |
| 1 | RVESV | both | rs9442216 | 1 | 16353400 | C | T | 1 | 0.67 | 0.04 | 0.01 | 18.6 | 1.60E-05 | 0.67 | 0.05 | 0.011 | 23.9 | 1.00E-06 | 0.67 | 0.03 | 0.006 | 36.8 | 1.30E-09 | 0.3 | 0.5598 | 0 | 0.5598 |
| 2 | RVESV | both | rs78529941 | 1 | 228551488 | A | G | 1 | 0.38 | 0.04 | 0.01 | 17.4 | 3.00E-05 | 0.38 | 0.06 | 0.01 | 37.1 | 1.10E-09 | 0.38 | 0.04 | 0.005 | 48.7 | 2.90E-12 | 2.9 | 0.0901 | 0.65 | 0.0901 |
| 2 | RVEF | both | rs78529941 | 1 | 228551488 | A | G | 1 | 0.38 | -0.03 | 0.011 | 9.7 | 1.80E-03 | 0.38 | -0.06 | 0.011 | 30.2 | 3.80E-08 | 0.38 | -0.04 | 0.007 | 35.8 | 2.20E-09 | 3.5 | 0.0622 | 0.71 | 0.0623 |
| 3 | RVESV | both | rs1314982 | 2 | 26922062 | A | G | 0.98 | 0.74 | -0.05 | 0.011 | 24.1 | 9.00E-07 | 0.74 | -0.04 | 0.012 | 13.6 | 2.30E-04 | 0.74 | -0.03 | 0.006 | 34.6 | 4.10E-09 | 0.4 | 0.5247 | 0 | 0.5247 |
| 3 | RVEDV | both | rs1731248 | 2 | 26924823 | C | A | 1 | 0.74 | -0.04 | 0.01 | 18.5 | 1.70E-05 | 0.75 | -0.04 | 0.011 | 11.5 | 7.00E-04 | 0.74 | -0.03 | 0.006 | 30 | 4.40E-08 | 0.1 | 0.7563 | 0 | 0.7563 |
| 4 | RVEDV | both | rs777900475 | 2 | 179477332 | A | AT | 0.99 | 0.21 | -0.04 | 0.011 | 13 | 3.10E-04 | 0.21 | -0.06 | 0.012 | 28.1 | 1.20E-07 | 0.21 | -0.04 | 0.006 | 41.3 | 1.30E-10 | 2.2 | 0.1343 | 0.55 | 0.1343 |
| 4 | RVESV | both | rs2042995 | 2 | 179558366 | C | T | 1 | 0.22 | -0.06 | 0.011 | 30.3 | 3.70E-08 | 0.22 | -0.07 | 0.012 | 37.6 | 8.80E-10 | 0.22 | -0.05 | 0.006 | 67.5 | 2.10E-16 | 0.7 | 0.4039 | 0 | 0.4039 |
| 4 | RVSV | both | rs7573293 | 2 | 179753245 | T | C | 0.99 | 0.73 | 0.04 | 0.011 | 15.3 | 9.40E-05 | 0.74 | 0.04 | 0.012 | 13.8 | 2.00E-04 | 0.73 | 0.04 | 0.007 | 32.5 | 1.20E-08 | 0 | 0.9433 | 0 | 0.9432 |
| 4 | RVEDV | both | rs955738 | 2 | 179775152 | C | T | 1 | 0.49 | 0.04 | 0.009 | 26 | 3.40E-07 | 0.49 | 0.05 | 0.01 | 22.5 | 2.10E-06 | 0.49 | 0.04 | 0.005 | 50.9 | 9.60E-13 | 0 | 0.8821 | 0 | 0.8821 |
| 5 | RVEF | men | rs55834511 | 3 | 14273414 | C | G | 0.99 | 0.22 | -0.04 | 0.013 | 8.7 | 3.20E-03 | 0.21 | -0.1 | 0.013 | 56.1 | 6.80E-14 | 0.22 | -0.06 | 0.009 | 53.1 | 3.20E-13 | 11.6 | 0.0006 | 0.91 | 0.0007 |
| 5 | RVESV | both | rs12630973 | 3 | 14293372 | T | G | 0.99 | 0.34 | -0.06 | 0.01 | 34.6 | 4.10E-09 | 0.35 | -0.06 | 0.011 | 30.5 | 3.30E-08 | 0.34 | -0.05 | 0.006 | 67.9 | 1.70E-16 | 0 | 0.8956 | 0 | 0.8955 |
| 6 | RVEDV | both | rs9832134 | 3 | 99836722 | C | T | 0.99 | 0.39 | -0.04 | 0.009 | 18.4 | 1.80E-05 | 0.39 | -0.04 | 0.01 | 12.8 | 3.40E-04 | 0.39 | -0.03 | 0.005 | 33.5 | 7.00E-09 | 0 | 0.8575 | 0 | 0.8575 |
| 7 | RVEF | women | rs9865460 | 3 | 158212019 | A | G | 0.99 | 0.47 | -0.06 | 0.01 | 36.9 | 1.20E-09 | 0.47 | -0.01 | 0.011 | 1.3 | 2.50E-01 | 0.47 | -0.04 | 0.007 | 28 | 1.20E-07 | 11.1 | 0.0008 | 0.91 | 0.0008 |
| 7 | RVESV | both | rs3851363 | 3 | 158328966 | A | C | 0.99 | 0.52 | 0.06 | 0.009 | 40 | 2.50E-10 | 0.52 | 0.05 | 0.01 | 20.1 | 7.20E-06 | 0.52 | 0.04 | 0.005 | 58.4 | 2.10E-14 | 0.9 | 0.3404 | 0 | 0.3404 |
| 7 | RVEDV | both | rs199736122 | 3 | 158423234 | CT | C | 0.98 | 0.16 | 0.05 | 0.012 | 19.4 | 1.00E-05 | 0.16 | 0.05 | 0.013 | 16.4 | 5.20E-05 | 0.16 | 0.04 | 0.007 | 34.4 | 4.40E-09 | 0 | 0.9696 | 0 | 0.9696 |
| 8 | RVEDV | both | rs200290730 | 4 | 120287492 | A | C | 0.98 | 0.32 | 0.04 | 0.01 | 19.3 | 1.10E-05 | 0.32 | 0.04 | 0.011 | 10.5 | 1.20E-03 | 0.32 | 0.03 | 0.006 | 29.8 | 4.70E-08 | 0.2 | 0.6326 | 0 | 0.6326 |
| 8 | RVESV | both | rs57023183 | 4 | 120636569 | GA | G | 0.99 | 0.64 | -0.04 | 0.01 | 20.7 | 5.40E-06 | 0.65 | -0.04 | 0.011 | 13.2 | 2.80E-04 | 0.64 | -0.03 | 0.006 | 36.9 | 1.20E-09 | 0.1 | 0.8163 | 0 | 0.8163 |
| 9 | RVSV | both | rs72967533 | 6 | 118655020 | C | T | 0.99 | 0.49 | 0.03 | 0.009 | 11.1 | 8.60E-04 | 0.48 | 0.05 | 0.01 | 22.4 | 2.30E-06 | 0.48 | 0.04 | 0.006 | 35.9 | 2.00E-09 | 1.6 | 0.2103 | 0.36 | 0.2102 |
| 9 | RVEDV | both | rs72967533 | 6 | 118655020 | C | T | 0.99 | 0.49 | 0.03 | 0.009 | 11.6 | 6.70E-04 | 0.48 | 0.04 | 0.01 | 17.5 | 2.80E-05 | 0.48 | 0.03 | 0.005 | 31.1 | 2.50E-08 | 0.6 | 0.4204 | 0 | 0.4204 |
| 10 | RVSV | men | rs3918226 | 7 | 150690176 | T | C | 0.97 | 0.08 | -0.03 | 0.017 | 3.4 | 6.60E-02 | 0.08 | -0.12 | 0.019 | 37.1 | 1.10E-09 | 0.08 | -0.06 | 0.011 | 32 | 1.60E-08 | 10.8 | 0.001 | 0.91 | 0.001 |
| 10 | RVEDV | men | rs3918226 | 7 | 150690176 | T | C | 0.97 | 0.08 | -0.04 | 0.016 | 4.7 | 3.10E-02 | 0.08 | -0.11 | 0.018 | 35.9 | 2.10E-09 | 0.08 | -0.05 | 0.01 | 31.7 | 1.80E-08 | 8.8 | 0.0029 | 0.89 | 0.0029 |
| 11 | RVEF | both | rs13274269 | 8 | 11449325 | T | G | 0.98 | 0.35 | 0.04 | 0.011 | 15.7 | 7.30E-05 | 0.34 | 0.03 | 0.015 | 4.5 | 3.30E-02 | 0.34 | 0.04 | 0.008 | 33.7 | 6.50E-09 | 0.4 | 0.5469 | 0 | 0.5469 |
| 12 | RVESV | both | 8:125858538 | 8 | 125858538 | G | GA | 0.98 | 0.31 | -0.04 | 0.01 | 20 | 7.70E-06 | 0.32 | -0.04 | 0.011 | 14.1 | 1.70E-04 | 0.31 | -0.03 | 0.006 | 30.8 | 2.90E-08 | 0 | 0.8815 | 0 | 0.8814 |
| 13 | RVESV | both | rs11784619 | 8 | 145013775 | A | G | 0.96 | 0.06 | -0.08 | 0.02 | 14.8 | 1.20E-04 | 0.06 | -0.1 | 0.022 | 22.4 | 2.20E-06 | 0.06 | -0.07 | 0.011 | 40.7 | 1.70E-10 | 0.8 | 0.3609 | 0 | 0.3609 |
| 13 | RVEF | both | rs11786896 | 8 | 145018354 | T | C | 0.98 | 0.05 | 0.1 | 0.024 | 18.7 | 1.60E-05 | 0.05 | 0.12 | 0.025 | 21.8 | 3.00E-06 | 0.05 | 0.11 | 0.016 | 40.9 | 1.60E-10 | 0.2 | 0.6843 | 0 | 0.6843 |
| 14 | RVESV | women | rs9298673 | 9 | 1224352 | C | A | 0.97 | 0.77 | 0.06 | 0.011 | 31.5 | 2.00E-08 | 0.77 | 0.01 | 0.012 | 1.2 | 2.80E-01 | 0.77 | 0.03 | 0.006 | 23.8 | 1.10E-06 | 9 | 0.0027 | 0.89 | 0.0027 |
| 15 | RVEF | both | rs2789750 | 9 | 35683473 | G | C | 0.99 | 0.32 | -0.06 | 0.011 | 29.7 | 5.10E-08 | 0.32 | -0.04 | 0.012 | 12.5 | 4.00E-04 | 0.32 | -0.05 | 0.008 | 41.2 | 1.40E-10 | 1.5 | 0.2271 | 0.31 | 0.2271 |
| 16 | RVEF | both | rs12006440 | 9 | 107703337 | T | C | 0.98 | 0.04 | -0.1 | 0.029 | 13.4 | 2.50E-04 | 0.03 | -0.12 | 0.031 | 16 | 6.20E-05 | 0.03 | -0.11 | 0.02 | 30 | 4.30E-08 | 0.2 | 0.622 | 0 | 0.622 |

| | | | | | | | | | | | | | | | | | | | | | | | | | | | |
|---|---|---|---|---|---|---|---|---|---|---|---|---|---|---|---|---|---|---|---|---|---|---|---|---|---|---|---|
| 17 | RVEDV | both | rs2066332 | 10 | 30322865 | G | A | 0.99 | 0.37 | -0.03 | 0.009 | 12.5 | 4.10E-04 | 0.37 | -0.05 | 0.01 | 22.6 | 2.00E-06 | 0.37 | -0.03 | 0.005 | 35.9 | 2.10E-09 | 1.4 | 0.2355 | 0.29 | 0.2355 |
| 18 | RVEF | women | rs111336312 | 10 | 88547019 | GA | G | 0.99 | 0.08 | 0.11 | 0.019 | 29.9 | 4.70E-08 | 0.08 | 0.01 | 0.02 | 0.6 | 4.50E-01 | 0.08 | 0.06 | 0.013 | 18.6 | 1.70E-05 | 10.6 | 0.0011 | 0.91 | 0.0011 |
| 19 | RVEF | both | rs72840788 | 10 | 121415685 | A | G | 0.98 | 0.22 | 0.08 | 0.013 | 40.5 | 1.90E-10 | 0.22 | 0.09 | 0.013 | 44.9 | 2.10E-11 | 0.22 | 0.08 | 0.009 | 87.5 | 8.30E-21 | 0.2 | 0.6231 | 0 | 0.6232 |
| 19 | RVESV | both | rs72840788 | 10 | 121415685 | A | G | 0.98 | 0.22 | -0.06 | 0.011 | 30.6 | 3.20E-08 | 0.22 | -0.08 | 0.012 | 47.2 | 6.40E-12 | 0.22 | -0.06 | 0.006 | 82.1 | 1.30E-19 | 1.5 | 0.2248 | 0.32 | 0.2248 |
| 20 | RVESV | both | rs3184504 | 12 | 111884608 | C | T | 1 | 0.52 | 0.06 | 0.009 | 38 | 7.10E-10 | 0.52 | 0.07 | 0.01 | 44.8 | 2.20E-11 | 0.52 | 0.05 | 0.005 | 80.4 | 3.10E-19 | 0.4 | 0.5521 | 0 | 0.5521 |
| 20 | RVSV | both | rs653178 | 12 | 112007756 | T | C | 1 | 0.52 | 0.04 | 0.009 | 18.6 | 1.60E-05 | 0.52 | 0.04 | 0.01 | 14.8 | 1.20E-04 | 0.52 | 0.03 | 0.006 | 30.3 | 3.70E-08 | 0 | 0.9939 | 0 | 0.9939 |
| 20 | RVEDV | both | rs653178 | 12 | 112007756 | T | C | 1 | 0.52 | 0.06 | 0.009 | 42.6 | 6.70E-11 | 0.52 | 0.06 | 0.01 | 39.1 | 4.10E-10 | 0.52 | 0.05 | 0.005 | 78.7 | 7.20E-19 | 0.1 | 0.7583 | 0 | 0.7583 |
| 21 | RVEDV | both | rs3825215 | 12 | 114804898 | C | G | 1 | 0.76 | -0.04 | 0.01 | 13.8 | 2.00E-04 | 0.75 | -0.05 | 0.011 | 20.2 | 7.10E-06 | 0.75 | -0.04 | 0.006 | 34.5 | 4.20E-09 | 0.6 | 0.4328 | 0 | 0.4328 |
| 21 | RVESV | both | rs1895606 | 12 | 114833384 | T | C | 0.98 | 0.46 | 0.04 | 0.009 | 19.9 | 8.20E-06 | 0.47 | 0.03 | 0.01 | 11.1 | 8.80E-04 | 0.47 | 0.03 | 0.005 | 32.3 | 1.30E-08 | 0.2 | 0.6308 | 0 | 0.6308 |
| 22 | RVSV | both | rs422068 | 14 | 23864804 | C | T | 1 | 0.36 | -0.05 | 0.01 | 27.1 | 2.00E-07 | 0.36 | -0.04 | 0.011 | 17.1 | 3.50E-05 | 0.36 | -0.04 | 0.006 | 42.8 | 6.10E-11 | 0.2 | 0.6765 | 0 | 0.6765 |
| 22 | RVEF | both | rs422068 | 14 | 23864804 | C | T | 1 | 0.36 | -0.05 | 0.011 | 20.3 | 6.70E-06 | 0.36 | -0.04 | 0.012 | 13.7 | 2.10E-04 | 0.36 | -0.04 | 0.007 | 32.8 | 1.00E-08 | 0.2 | 0.6754 | 0 | 0.6754 |
| 23 | RVEDV | both | rs62048478 | 16 | 53429180 | T | C | 0.99 | 0.32 | 0.04 | 0.009 | 15.2 | 9.80E-05 | 0.32 | 0.04 | 0.011 | 16 | 6.50E-05 | 0.32 | 0.03 | 0.006 | 33.1 | 8.70E-09 | 0.2 | 0.6717 | 0 | 0.6717 |
| 23 | RVESV | both | rs113335556 | 16 | 53514820 | T | C | 1 | 0.3 | 0.04 | 0.01 | 14.8 | 1.20E-04 | 0.3 | 0.04 | 0.011 | 11.3 | 7.70E-04 | 0.3 | 0.03 | 0.006 | 30.6 | 3.30E-08 | 0 | 0.8823 | 0 | 0.8823 |
| 24 | RVESV | both | rs17608766 | 17 | 45013271 | C | T | 1 | 0.15 | 0.07 | 0.013 | 29.6 | 5.40E-08 | 0.15 | 0.07 | 0.014 | 26 | 3.40E-07 | 0.15 | 0.06 | 0.007 | 53.3 | 2.90E-13 | 0 | 0.9987 | 0 | 0.9987 |
| 24 | RVEDV | both | rs76774446 | 17 | 45046368 | A | C | 1 | 0.14 | 0.07 | 0.013 | 25.4 | 4.50E-07 | 0.14 | 0.07 | 0.014 | 23.2 | 1.50E-06 | 0.14 | 0.05 | 0.007 | 47.8 | 4.80E-12 | 0 | 0.8999 | 0 | 0.8999 |
| 25 | RVEF | both | 18:34235735 | 18 | 34235735 | C | CAA | 0.98 | 0.25 | 0.05 | 0.012 | 19.4 | 1.00E-05 | 0.25 | 0.06 | 0.013 | 25.1 | 5.30E-07 | 0.25 | 0.05 | 0.008 | 41.2 | 1.40E-10 | 0.4 | 0.5205 | 0 | 0.5205 |
| 26 | RVESV | both | rs7251903 | 19 | 39184811 | A | G | 1 | 0.49 | 0.05 | 0.009 | 26.1 | 3.30E-07 | 0.48 | 0.03 | 0.01 | 11.5 | 7.10E-04 | 0.48 | 0.03 | 0.005 | 34.9 | 3.50E-09 | 1 | 0.3094 | 0.03 | 0.3094 |
| 27 | RVESV | both | rs56096557 | 19 | 46310895 | C | A | 1 | 0.35 | 0.04 | 0.01 | 17.6 | 2.70E-05 | 0.35 | 0.06 | 0.011 | 28.3 | 1.00E-07 | 0.35 | 0.04 | 0.006 | 46.3 | 9.90E-12 | 1.2 | 0.2782 | 0.15 | 0.2782 |
| 28 | RVSV | both | rs4811602 | 20 | 36849088 | A | G | 0.98 | 0.47 | 0.05 | 0.009 | 29.5 | 5.60E-08 | 0.47 | 0.03 | 0.01 | 8.5 | 3.50E-03 | 0.47 | 0.04 | 0.006 | 33.7 | 6.30E-09 | 2.3 | 0.1318 | 0.56 | 0.1318 |

Data are sorted by chromosome (CHR) and base pair (BP). Loci are labelled randomly from 1 to 28.

* Sex with most significant association (lowest P-value) is given.

Chi-Square (CHISQ). Effect allele (EA). Effect allele frequency (EAF). Other allele (OA). RV end-diastolic volume (RVEDV). RV end-diastolic volume (RVEDV). RV end-systolic volume (RVESV). RV stroke volume (RVSV). RV ejection fraction (RVEF). Standard error (SE).

**Table S5:** Replication of association pairs in sex-combined analyses in the identical population.

| Locus, phenotype, variant | | | | | | | | Aung et al (PMID: 35697868) | | | | | | Pirruccello et al (PMID: 35697867) | | | |
|---|---|---|---|---|---|---|---|---|---|---|---|---|---|---|---|---|---|
| Locus | Phenotype | RSID | CHR | BP (GRCh37) | EA | OA | EAF[*] | LD proxy | EAF | Beta | SE | pval | LD proxy | EAF | Beta | SE | P |
| 1 | RVEF | rs9442214 | 1 | 16349460 | C | T | 0.67 | No | 0.33 | -0.05 | 0.009 | 5.10E-08 | No | 0.33 | -0.04 | 0.007 | 7.10E-10 |
| 1 | RVESV | rs9442216 | 1 | 16353400 | C | T | 0.67 | No | 0.33 | 0.05 | 0.008 | 5.10E-08 | No | 0.33 | 0.03 | 0.006 | 1.50E-07 |
| 2 | RVESV | rs78529941 | 1 | 228551488 | A | G | 0.38 | No | 0.62 | 0.06 | 0.008 | 7.10E-13 | No | 0.62 | 0.03 | 0.006 | 1.60E-08 |
| 2 | RVEF | rs78529941 | 1 | 228551488 | A | G | 0.38 | No | 0.62 | -0.05 | 0.008 | 3.50E-10 | No | 0.62 | -0.03 | 0.007 | 4.60E-06 |
| 3 | RVESV | rs1314982 | 2 | 26922062 | A | G | 0.74 | No | 0.26 | -0.04 | 0.009 | 2.50E-05 | No | 0.26 | -0.04 | 0.006 | 3.20E-10 |
| 3 | RVEDV | rs1731248 | 2 | 26924823 | C | A | 0.74 | No | 0.26 | -0.03 | 0.009 | 6.70E-04 | No | 0.26 | -0.03 | 0.006 | 2.00E-06 |
| 4 | RVEDV | rs777900475 | 2 | 179477332 | A | AT | 0.21 | No | 0.79 | -0.07 | 0.01 | 2.50E-12 | No | 0.78 | -0.02 | 0.006 | 1.40E-03 |
| 4 | RVESV | rs2042995 | 2 | 179558366 | C | T | 0.22 | No | 0.78 | -0.08 | 0.01 | 4.00E-18 | No | 0.77 | -0.04 | 0.006 | 6.30E-09 |
| 4 | RVSV | rs7573293 | 2 | 179753245 | T | C | 0.73 | No | 0.27 | 0.04 | 0.009 | 4.50E-06 | No | 0.28 | 0.03 | 0.007 | 4.20E-06 |
| 4 | RVEDV | rs955738 | 2 | 179775152 | C | T | 0.49 | No | 0.51 | 0.04 | 0.008 | 1.70E-07 | No | 0.52 | 0.03 | 0.005 | 2.20E-07 |
| 5 | RVEF | rs55834511 | 3 | 14273414 | C | G | 0.22 | No | 0.79 | -0.07 | 0.01 | 5.30E-13 | No | 0.79 | -0.05 | 0.008 | 7.00E-10 |
| 5 | RVESV | rs12630973 | 3 | 14293372 | T | G | 0.34 | No | 0.66 | -0.07 | 0.009 | 5.60E-15 | No | 0.66 | -0.05 | 0.006 | 8.10E-16 |
| 6 | RVEDV | rs9832134 | 3 | 99836722 | C | T | 0.39 | No | 0.61 | -0.04 | 0.008 | 1.40E-07 | No | 0.61 | -0.03 | 0.005 | 3.80E-07 |
| 7 | RVEF | rs9865460 | 3 | 158212019 | A | G | 0.47 | No | 0.53 | -0.04 | 0.008 | 7.60E-08 | No | 0.53 | -0.04 | 0.007 | 3.60E-10 |
| 7 | RVESV | rs3851363 | 3 | 158328966 | A | C | 0.52 | No | 0.48 | 0.06 | 0.008 | 5.80E-12 | No | 0.48 | 0.04 | 0.005 | 1.30E-10 |
| 7 | RVEDV | rs199736122 | 3 | 158423234 | CT | C | 0.16 | No | 0.84 | 0.05 | 0.011 | 1.00E-06 | No | 0.84 | 0.03 | 0.007 | 3.30E-05 |
| 8 | RVEDV | rs200290730 | 4 | 120287492 | A | C | 0.32 | No | 0.68 | 0.05 | 0.009 | 7.50E-08 | No | 0.68 | 0.02 | 0.006 | 2.90E-04 |
| 8 | RVESV | rs57023183 | 4 | 120636569 | GA | G | 0.64 | No | 0.36 | -0.05 | 0.008 | 3.20E-08 | No | 0.36 | -0.02 | 0.006 | 2.10E-03 |
| 9 | RVSV | rs72967533 | 6 | 118655020 | C | T | 0.48 | No | 0.52 | 0.04 | 0.008 | 6.90E-08 | No | 0.52 | 0.02 | 0.006 | 1.00E-03 |
| 9 | RVEDV | rs72967533 | 6 | 118655020 | C | T | 0.48 | No | 0.52 | 0.04 | 0.008 | 3.40E-07 | No | 0.52 | 0.02 | 0.005 | 4.30E-05 |
| 10 | RVSV | rs3918226 | 7 | 150690176 | T | C | 0.08 | No | 0.92 | -0.08 | 0.015 | 8.40E-08 | No | 0.92 | -0.05 | 0.011 | 2.60E-05 |
| 10 | RVEDV | rs3918226 | 7 | 150690176 | T | C | 0.08 | No | 0.92 | -0.08 | 0.015 | 1.50E-07 | No | 0.92 | -0.05 | 0.01 | 3.60E-07 |
| 11 | RVEF | rs13274269 | 8 | 11449325 | T | G | 0.34 | No | 0.66 | 0.04 | 0.009 | 9.80E-07 | No | 0.66 | 0.04 | 0.007 | 5.70E-09 |
| 12 | RVESV | 8:125858538 | 8 | 125858538 | G | GA | 0.31 | Yes[†] | 0.69 | -0.04 | 0.009 | 3.20E-06 | No | 0.69 | -0.03 | 0.006 | 4.40E-07 |
| 13 | RVESV | rs11784619 | 8 | 145013775 | A | G | 0.06 | No | 0.94 | -0.1 | 0.017 | 4.40E-08 | No | 0.94 | -0.07 | 0.012 | 6.00E-08 |
| 13 | RVEF | rs11786896 | 8 | 145018354 | T | C | 0.05 | No | 0.95 | 0.11 | 0.019 | 4.20E-09 | No | 0.95 | 0.09 | 0.016 | 3.60E-09 |
| 14 | RVESV | rs9298673 | 9 | 1224352 | C | A | 0.77 | No | 0.23 | 0.03 | 0.01 | 6.20E-04 | No | 0.23 | 0.02 | 0.007 | 1.20E-03 |
| 15 | RVEF | rs2789750 | 9 | 35683473 | G | C | 0.32 | No | 0.68 | -0.06 | 0.009 | 8.40E-11 | No | 0.68 | -0.04 | 0.007 | 1.40E-06 |

| | | | | | | | | | | | | | | | | | |
|---|---|---|---|---|---|---|---|---|---|---|---|---|---|---|---|---|---|
| 16 | RVEF | rs12006440 | 9 | 107703337 | T | C | 0.03 | No | 0.97 | -0.14 | 0.023 | 2.40E-09 | No | 0.97 | -0.09 | 0.019 | 9.80E-06 |
| 17 | RVEDV | rs2066332 | 10 | 30322865 | G | A | 0.37 | No | 0.63 | -0.05 | 0.008 | 4.10E-08 | No | 0.63 | -0.02 | 0.006 | 1.30E-05 |
| 18 | RVEF | rs111336312 | 10 | 88547019 | GA | G | 0.08 | No | 0.92 | 0.05 | 0.015 | 4.80E-04 | No | 0.92 | 0.02 | 0.013 | 1.20E-01 |
| 19 | RVEF | rs72840788 | 10 | 121415685 | A | G | 0.22 | No | 0.78 | 0.09 | 0.01 | 8.00E-19 | No | 0.78 | 0.08 | 0.008 | 8.40E-19 |
| 19 | RVESV | rs72840788 | 10 | 121415685 | A | G | 0.22 | No | 0.78 | -0.08 | 0.01 | 2.70E-17 | No | 0.78 | -0.06 | 0.007 | 3.80E-19 |
| 20 | RVESV | rs3184504 | 12 | 111884608 | C | T | 0.52 | No | 0.48 | 0.06 | 0.008 | 4.20E-14 | No | 0.47 | 0.05 | 0.005 | 4.40E-17 |
| 20 | RVSV | rs653178 | 12 | 112007756 | T | C | 0.52 | No | 0.48 | 0.04 | 0.008 | 3.50E-07 | No | 0.47 | 0.03 | 0.006 | 1.10E-08 |
| 20 | RVEDV | rs653178 | 12 | 112007756 | T | C | 0.52 | No | 0.48 | 0.06 | 0.008 | 1.70E-14 | No | 0.47 | 0.04 | 0.005 | 1.10E-16 |
| 21 | RVEDV | rs3825215 | 12 | 114804898 | C | G | 0.75 | No | 0.25 | -0.05 | 0.009 | 6.90E-08 | No | 0.25 | -0.02 | 0.006 | 6.80E-04 |
| 21 | RVESV | rs1895606 | 12 | 114833384 | T | C | 0.47 | No | 0.54 | 0.04 | 0.008 | 1.80E-07 | No | 0.54 | 0.02 | 0.005 | 4.60E-03 |
| 22 | RVSV | rs422068 | 14 | 23864804 | C | T | 0.36 | No | 0.64 | -0.05 | 0.009 | 1.10E-07 | No | 0.64 | -0.02 | 0.006 | 5.20E-05 |
| 22 | RVEF | rs422068 | 14 | 23864804 | C | T | 0.36 | No | 0.64 | -0.03 | 0.009 | 1.30E-04 | No | 0.64 | -0.01 | 0.007 | 3.20E-02 |
| 23 | RVEDV | rs62048478 | 16 | 53429180 | T | C | 0.32 | No | 0.68 | 0.05 | 0.009 | 1.90E-08 | No | 0.68 | 0.02 | 0.006 | 6.10E-05 |
| 23 | RVESV | rs113335556 | 16 | 53514820 | T | C | 0.3 | No | 0.7 | 0.05 | 0.009 | 2.20E-08 | No | 0.7 | 0.03 | 0.006 | 1.00E-05 |
| 24 | RVESV | rs17608766 | 17 | 45013271 | C | T | 0.15 | No | 0.85 | 0.08 | 0.011 | 6.10E-12 | No | 0.86 | 0.05 | 0.008 | 3.60E-09 |
| 24 | RVEDV | rs76774446 | 17 | 45046368 | A | C | 0.14 | No | 0.86 | 0.07 | 0.012 | 1.50E-08 | No | 0.87 | 0.05 | 0.008 | 7.00E-11 |
| 25 | RVEF | 18:34235735 | 18 | 34235735 | C | CAA | 0.25 | Yes‡ | 0.75 | 0.06 | 0.009 | 2.90E-10 | No | 0.75 | 0.04 | 0.008 | 1.90E-07 |
| 26 | RVESV | rs7251903 | 19 | 39184811 | A | G | 0.48 | No | 0.52 | 0.05 | 0.008 | 1.90E-09 | No | 0.52 | 0.02 | 0.005 | 4.10E-04 |
| 27 | RVESV | rs56096557 | 19 | 46310895 | C | A | 0.35 | No | 0.65 | 0.05 | 0.008 | 3.20E-09 | No | 0.65 | 0.03 | 0.006 | 2.60E-07 |
| 28 | RVSV | rs4811602 | 20 | 36849088 | A | G | 0.47 | No | 0.53 | 0.05 | 0.008 | 5.70E-08 | No | 0.53 | 0.03 | 0.006 | 1.20E-05 |

Data are sorted by chromosome (CHR) and base pair (BP). Loci are labelled randomly from 1 to 28.

* Effect allele frequency (EAF) from sex-combined data are given.

† A proxy rs72716972 in high linkage disequilibrium (LD) with the lead variant ($r^2$=0.99) were used.

‡ A proxy rs2644267 in high linkage disequilibrium (LD) with the lead variant ($r^2$=0.96) were used.

Effect allele (EA). Other allele (OA). RV end-diastolic volume (RVEDV). RV end-diastolic volume (RVEDV). RV end-systolic volume (RVESV). RV stroke volume (RVSV). RV ejection fraction (RVEF). Standard error (SE).

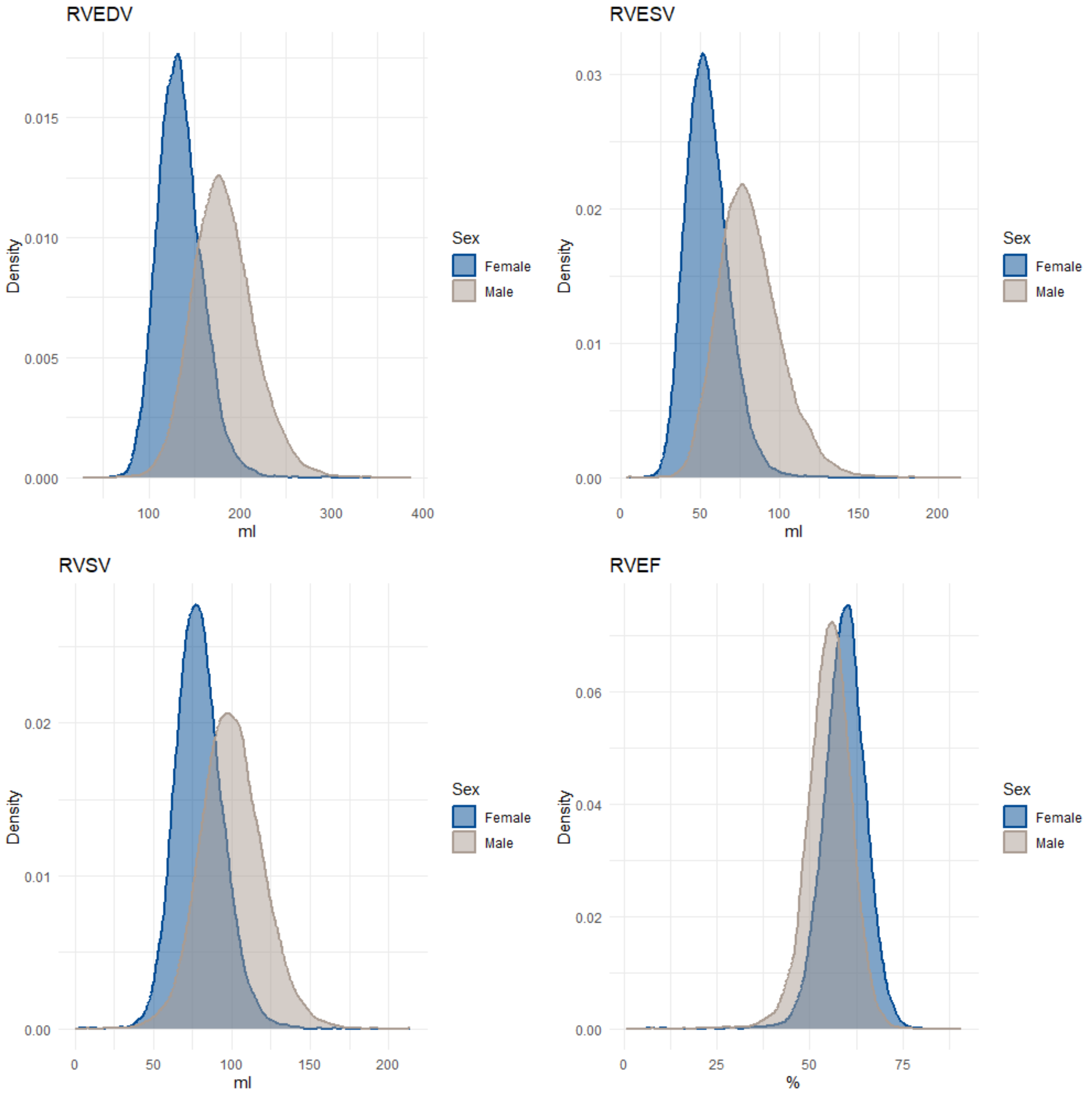


**Figure S1**. Density plots illustrating the distribution of right ventricular (RV) phenotypes by sex. The plots represent the probability density of the phenotypes, with kernel density estimation smoothing applied for clarity. The peak regions indicate areas of higher data concentration, while the tails represent lower density regions.

RV end-diastolic volume (RVEDV). RV end-systolic volume (RVESV). RV stroke volume (RVSV). RV ejection fraction (RVEF).

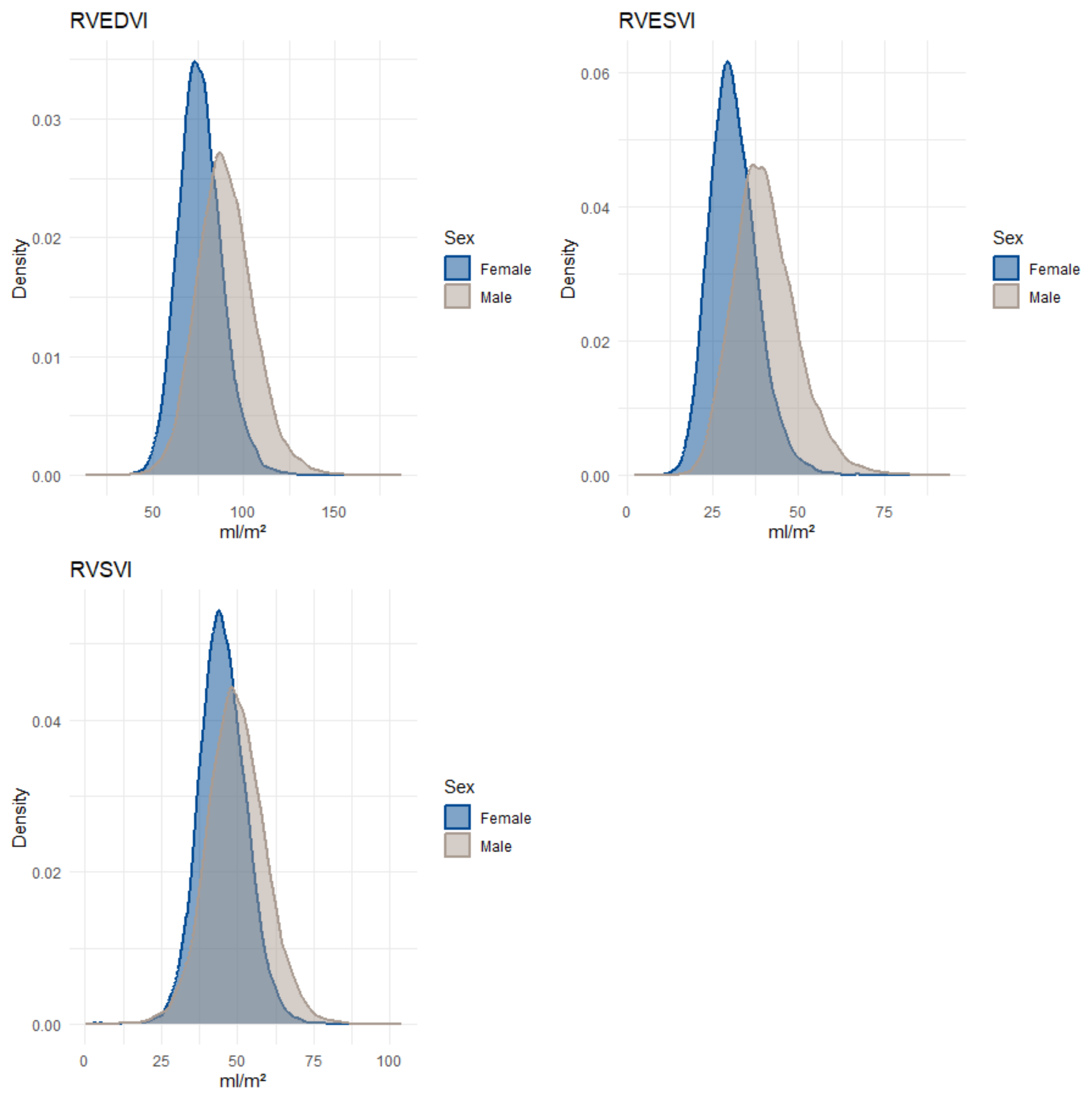


**Figure S2**. Density plots illustrating the distribution of right ventricular (RV) phenotypes by sex. The plots represent the probability density of the phenotypes, with kernel density estimation smoothing applied for clarity. The peak regions indicate areas of higher data concentration, while the tails represent lower density regions. Right ventricular volumes were indexed for body surface area.

Indexed RV end-diastolic volume (RVEDVI). Indexed RV end-systolic volume (RVESVI). Indexed RV stroke volume (RVSVI).

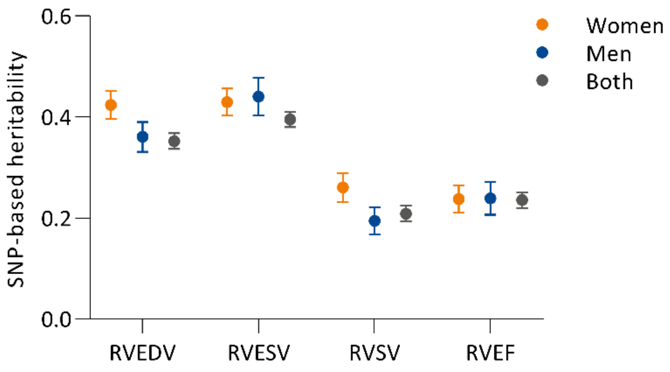


**Figure S3**: Sex difference in SNP-based heritability. Estimation of heritability with standard error included hard-called genotypes on all chromosomes combined (1-22 and X).

RV end-diastolic volume (RVEDV). RV end-systolic volume (RVESV). RV stroke volume (RVSV). RV ejection fraction (RVEF). Single-nucleotide polymorphism (SNP).

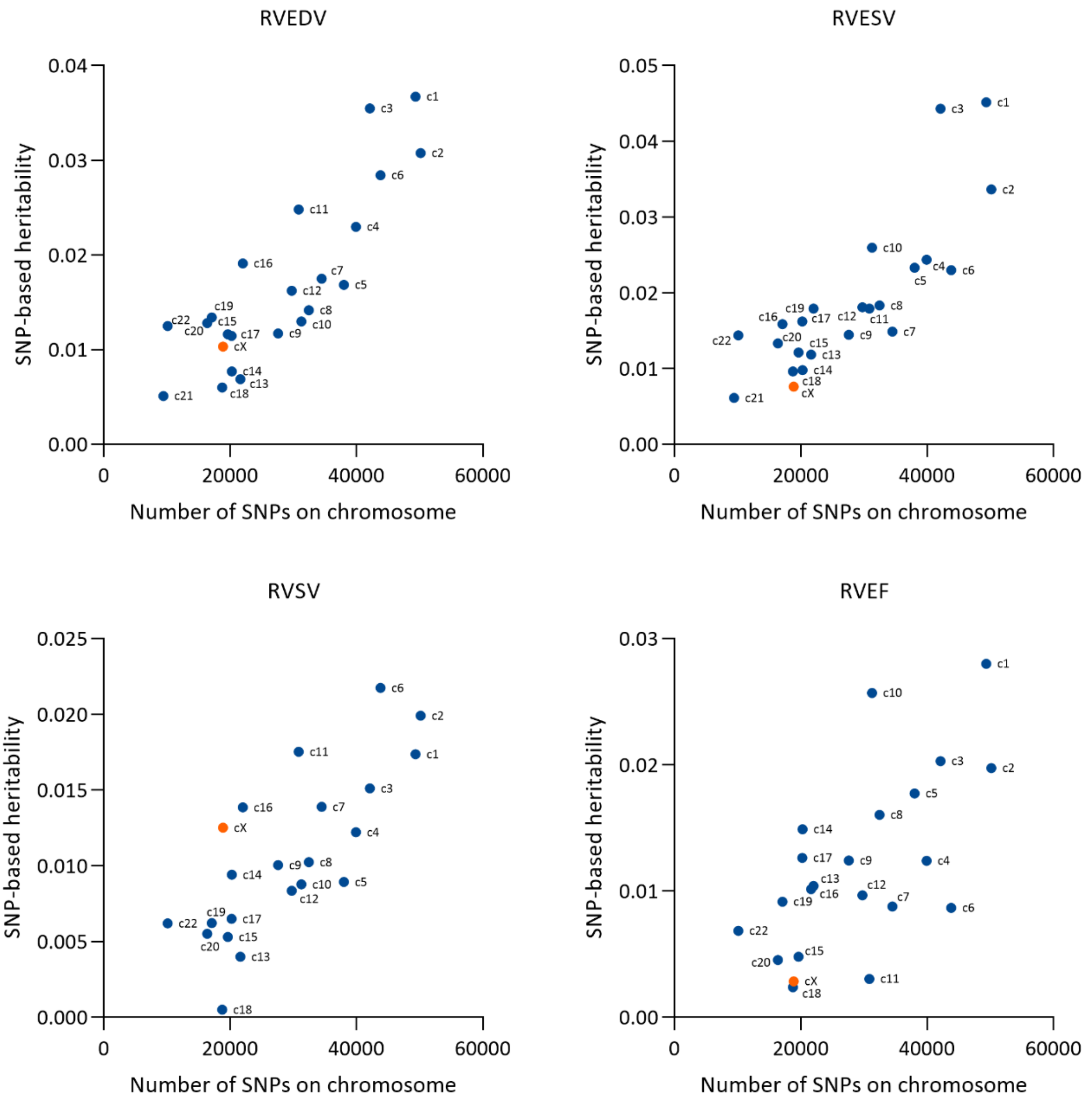


**Figure S4:** SNP-based estimation of heritability of RV traits by chromosome in the combined analyses of both sexes. Hard called genotypes were split into each individual chromosome and separate genetic relationship matrices were created. Heritability attributable to were estimated in a restricted maximum likelihood analysis including top genotype principal components and other covariates. Heritability for each chromosome were plotted against the total number of variants included per chromosome. The X-chromosome was highlighted in orange. Heritability estimates were <0.001 for chromosome 21 on RVSV and RVEF (not included).

RV end-diastolic volume (RVEDV). RV end-systolic volume (RVESV). RV stroke volume (RVSV). RV ejection fraction (RVEF). Single-nucleotide polymorphism (SNP).

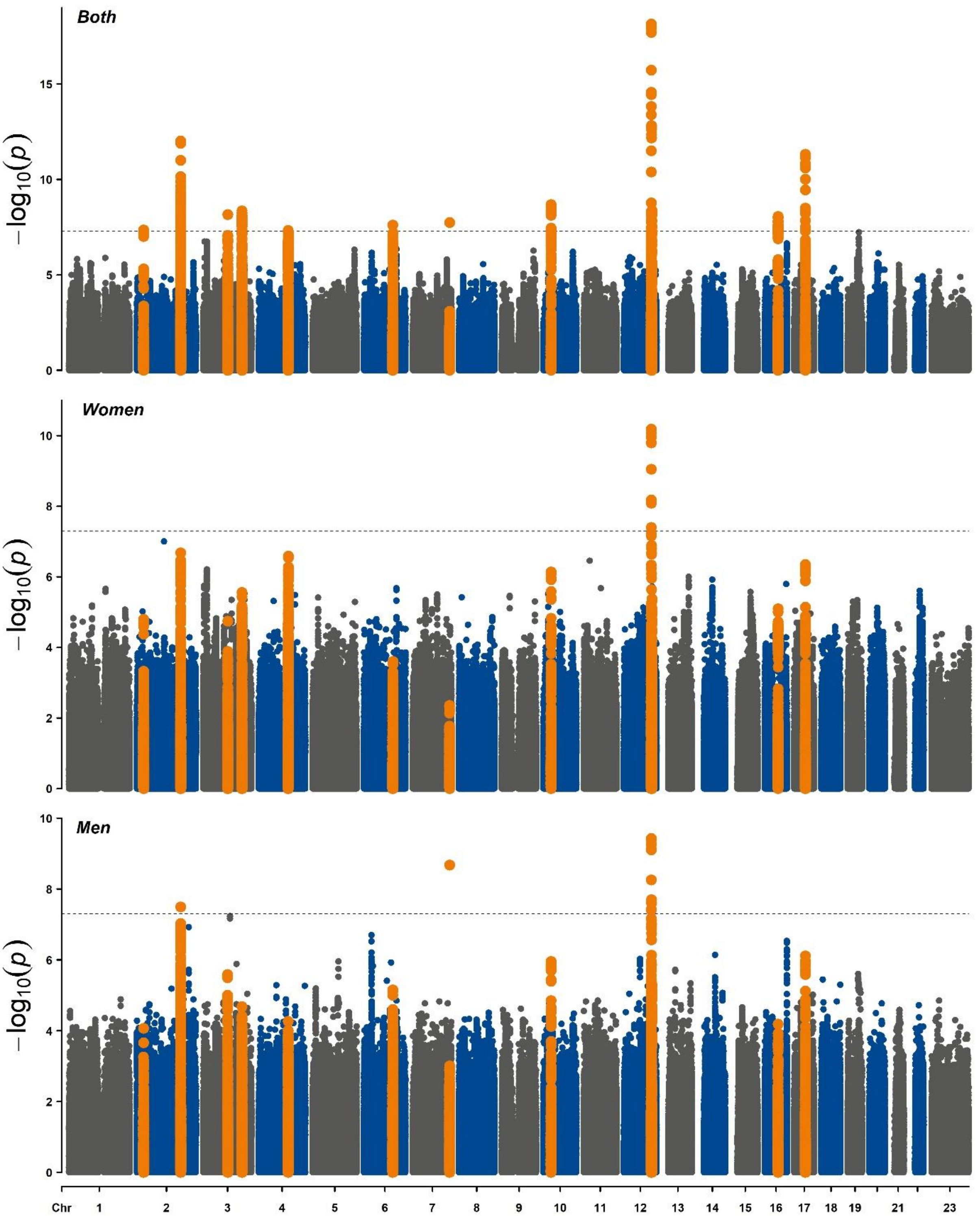


**Figure S5**: Manhattan plot for genome-wide association studies on right ventricular end-diastolic volume (RVEDV) in women, men and both sexes combined. Highlighted variants are those in linkage disequilibrium with the respective lead variant (LD>0.001).

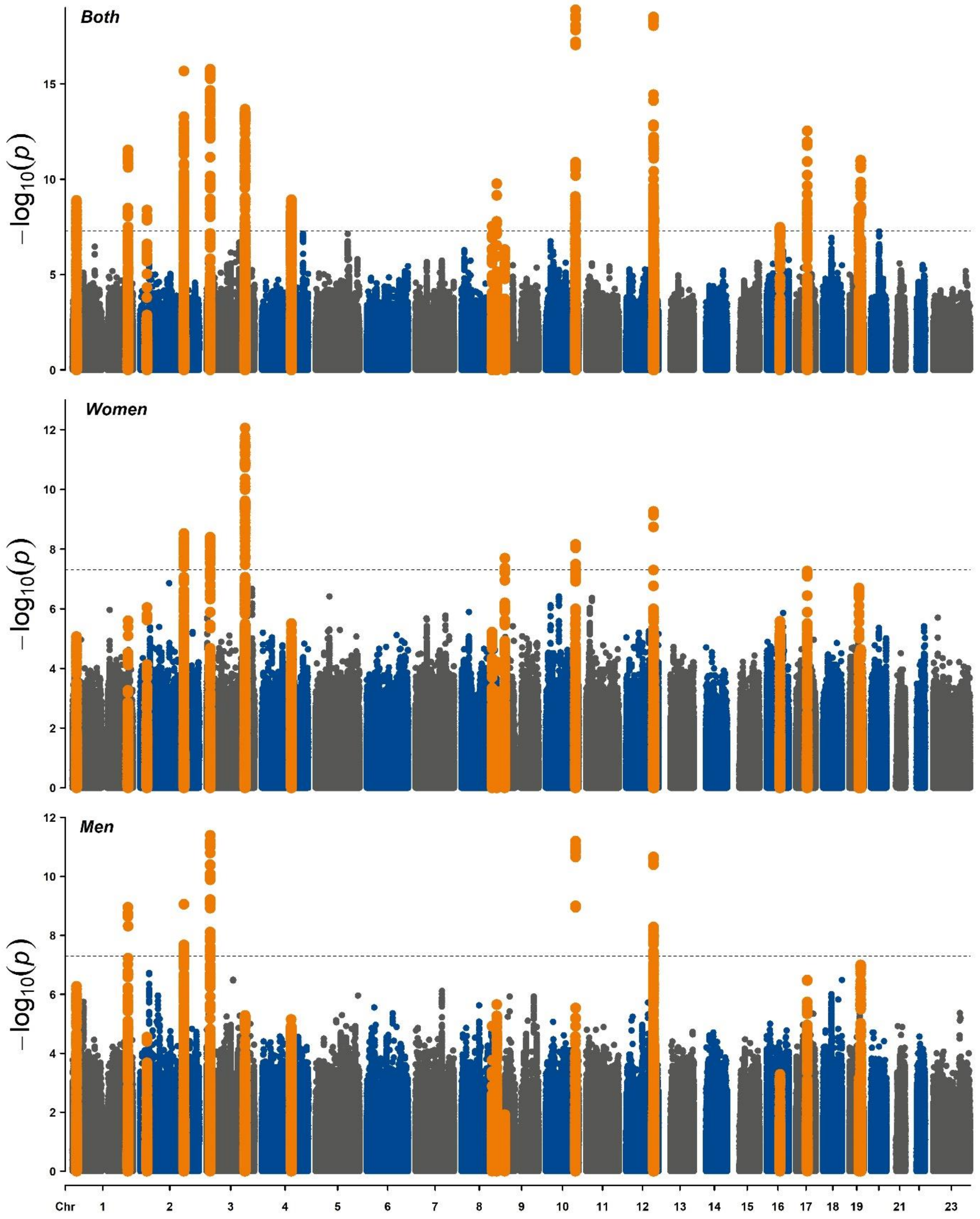


**Figure S6:** Manhattan plot for genome-wide association studies on right ventricular end-systolic volume (RVESV) in women, men and both sexes combined. Highlighted variants are those in linkage disequilibrium with the respective lead variant (LD>0.001).

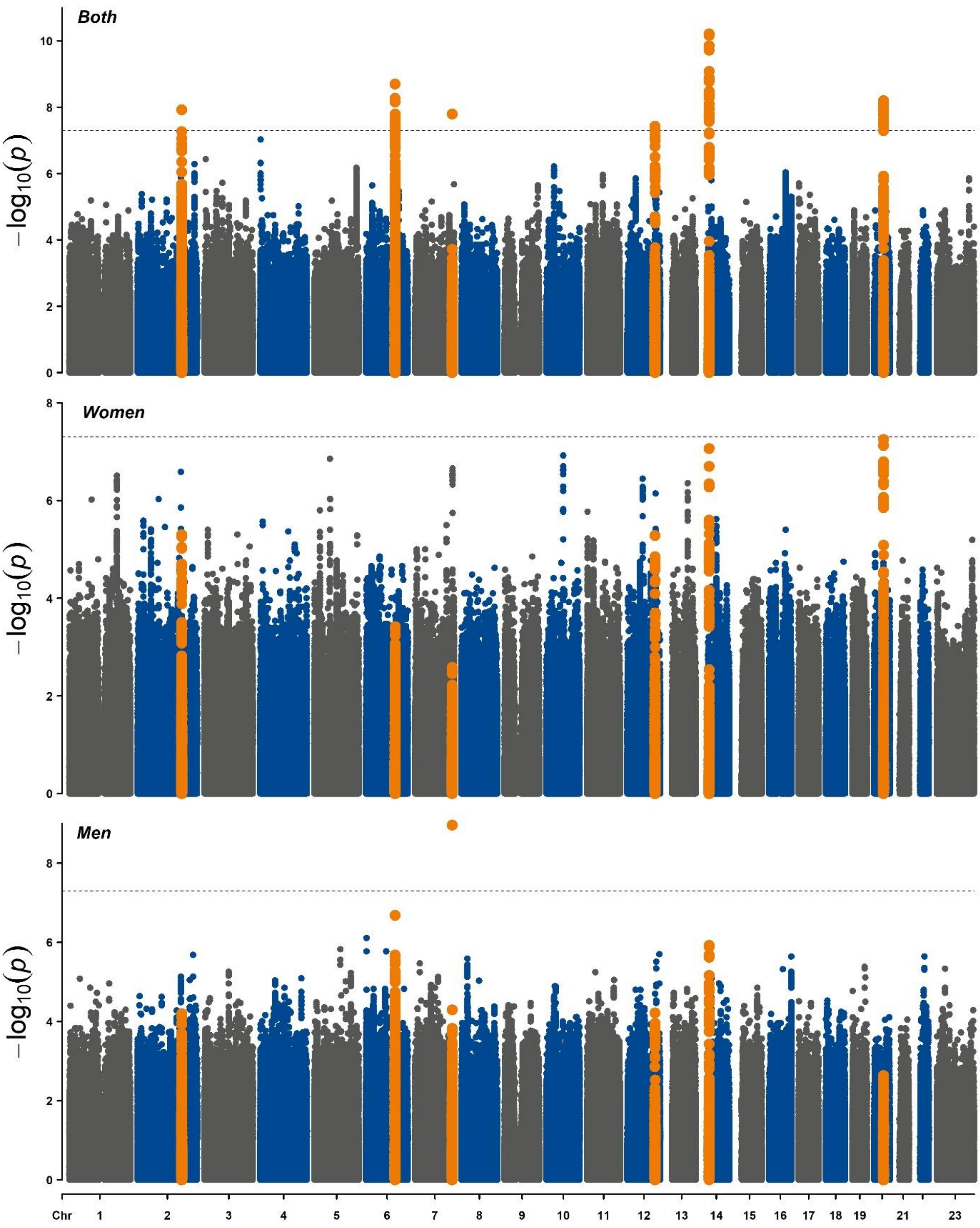


**Figure S7:** Manhattan plot for genome-wide association studies on right ventricular stroke volume (RVSV) in women, men and both sexes combined. Highlighted variants are those in linkage disequilibrium with the respective lead variant (LD>0.001).

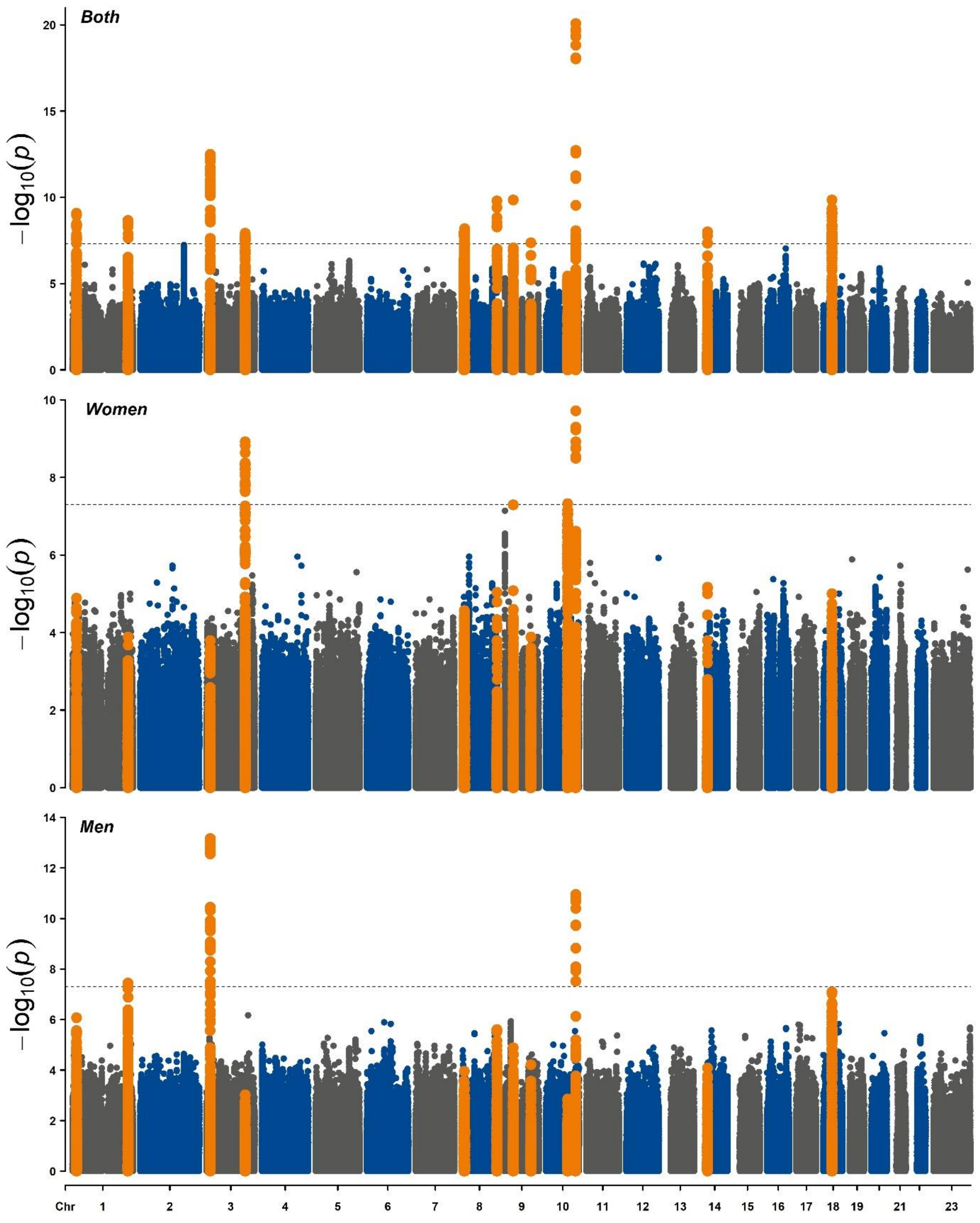


**Figure S8:** Manhattan plot for genome-wide association studies on right ventricular end-ejection fraction (RVEF) in women, men and both sexes combined. Highlighted variants are those in linkage disequilibrium with the respective lead variant (LD>0.001).

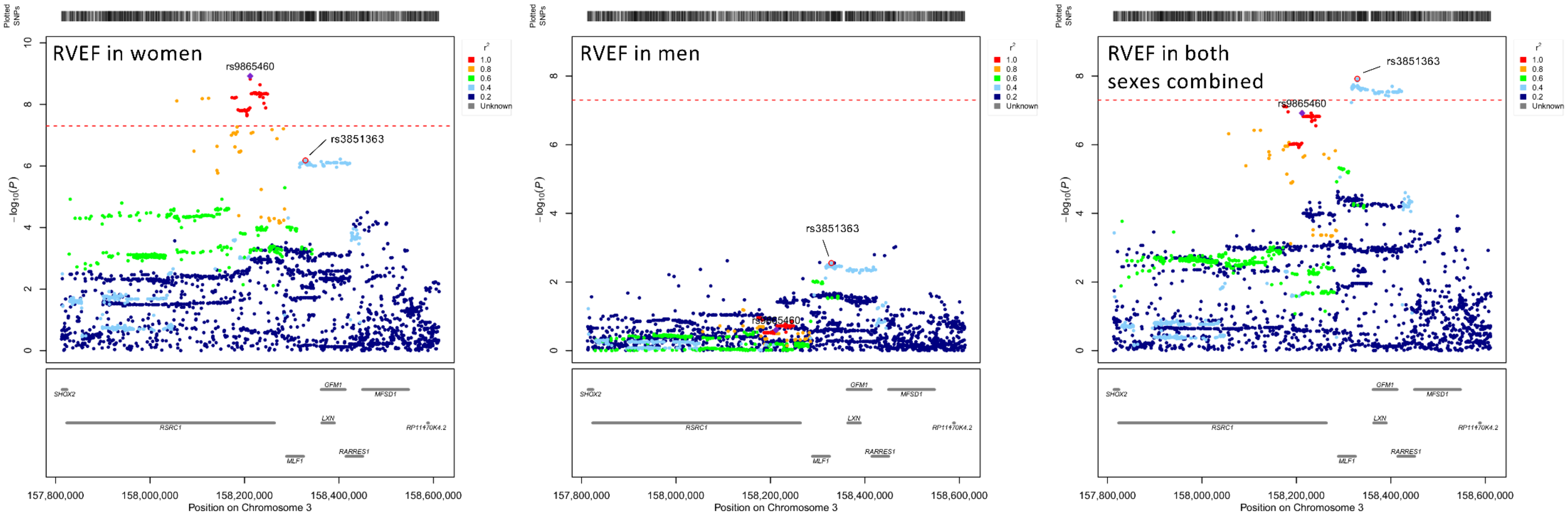


**Figure S9:** Regional association in women, men and both sexes combined for right ventricular ejection fraction (RVEF) at *RSRC1* locus. Regional plots show the genomic positions of variants on the x-axis and their association strength (-log10(p-value)) on the y-axis. Correlation coefficients ($r^2$) with the lead variant (rs9865460) are indicated by colour. Lead variants with the most significant p-values were different between women, men and the combined data. The lead variant in women (rs9865460) displayed the most significant p-value among all sexes and was found to be in linkage disequilibrium ($r^2$=0.36) with the lead variants observed in the combined sex analysis (rs3851363). Posterior probability for a shared causal variant (H4) between women and men was 1.7%

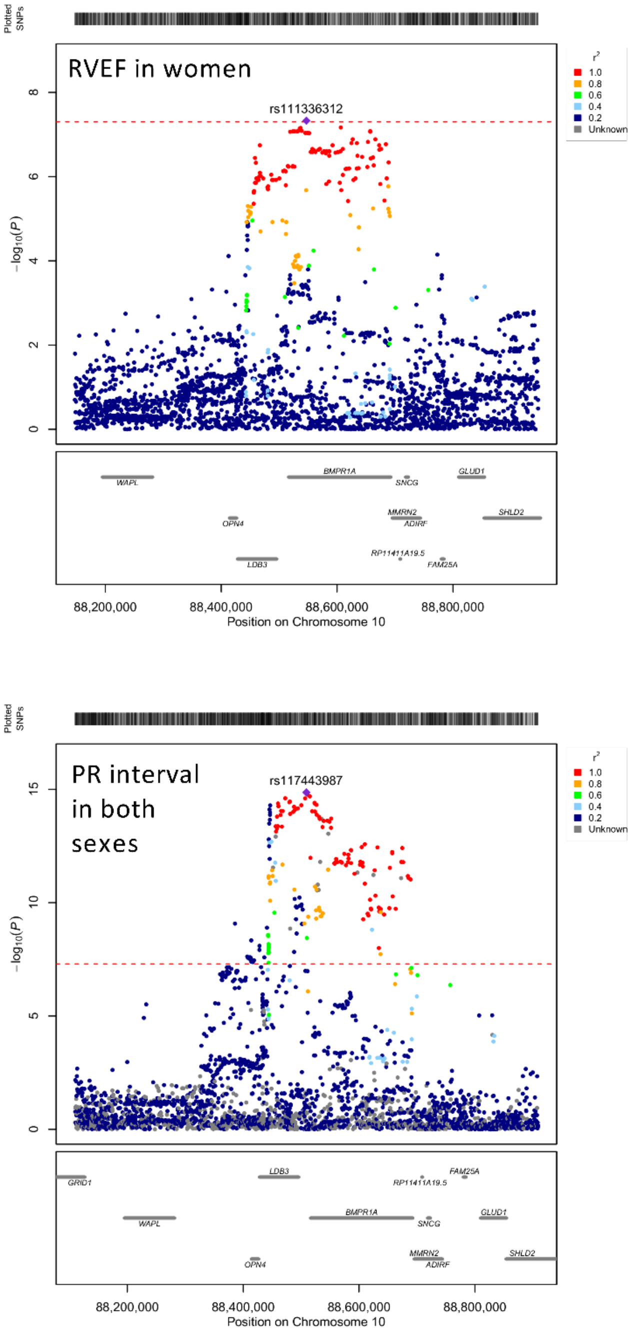


Ntalla et al. Nat Commun. 2020 May 21;11(1):2542.

**Figure S10:** Regional association at the *BMPR1A* locus for right ventricular ejection fraction (RVEF) in women and electrocardiographic PR interval in both sexes combined on European individuals. Plots show the genomic positions of variants on the x-axis and their association strength (-log10(p-value)) on the y-axis. Correlation coefficients ($r^2$) with the lead variant are indicated by colour. The lead variant for RVEF in women (rs111336312) was found to be in linkage disequilibrium with the lead variant for PR interval in both sexes (rs117443987; $r^2$=0.97). Posterior probability for a shared causal variant (H4) between the two associations was 86%.
